# Consortium Profile: The Methylation, Imaging and NeuroDevelopment (MIND) Consortium

**DOI:** 10.1101/2024.06.23.24309353

**Authors:** Isabel K. Schuurmans, Rosa H. Mulder, Vilte Baltramonaityte, Alexandra Lahtinen, Fan Qiuyu, Leonardo Melo Rothmann, Marlene Staginnus, Jetro Tuulari, S. Alexandra Burt, Claudia Buss, Jeffrey M. Craig, Kirsten A. Donald, Janine F. Felix, Tom P. Freeman, Rodrigo Grassi-Oliveira, Anke Huels, Luke W. Hyde, Scott A. Jones, Hasse Karlsson, Linnea Karlsson, Nastassja Koen, Will Lawn, Colter Mitchell, Christopher S. Monk, Michael A. Mooney, Ryan Muetzel, Joel T. Nigg, Síntia Iole Nogueira Belangero, Daniel Notterman, Tom O’Connor, Kieran J. O’Donnell, Pedro Mario Pan, Tiina Paunio, Peter Ryabinin, Richard Saffery, Giovanni A. Salum, Marc Seal, Tim J. Silk, Dan J. Stein, Heather Zar, Esther Walton, Charlotte A. M. Cecil

**Affiliations:** Department of Child and Adolescent Psychiatry and Psychology, Erasmus MC University Medical Center Rotterdam, Rotterdam, the Netherlands; Department of Psychology, University of Bath, Bath, United Kingdom; Research Program in Systems Oncology, Research Programs Unit, Faculty of Medicine, University of Helsinki; National institute of Health and welfare, Department of Public Health and Welfare, Population Health Unit; Translational Neuropsychiatry Unit, Department of Clinical Medicine, Aarhus University, Palle Juul-Jensens Boulevard 11, A701-129, 8200, Aarhus, Denmark; FinnBrain Birth Cohort Study, Turku Brain and Mind Center; Department of Clinical Medicine, University of Turku, Turku, Finland; Centre for Population Health Research, Turku University Hospital and University of Turku, Turku, Finland; Michigan State University; Development, Health and Disease Research Program, University of California, Irvine, California, USA.; Deakin University, IMPACT the Institute for Mental and Physical Health and Clinical Translation, School of Medicine, Geelong, Australia; Department of Paediatrics and Child Health, Red Cross War Memorial Childrens Hospital, University of Cape Town, Cape Town, South Africa; The Generation R Study Group, Erasmus MC University Medical Center Rotterdam, Rotterdam, the Netherlands; Department of Epidemiology, Rollins School of Public Health, Emory University, Atlanta, GA, USA; Department of Psychology, University of Michigan, Ann Arbor, Michigan, USA; Center for Mental Health Innovation, Oregon Health & Science University, Portland, Oregon, USA; Department of Psychiatry and Mental Health, University of Cape Town, Cape Town, South Africa; Department of Psychology, Institute of Psychiatry Psychology and Neuroscience, King's College London, London, UK.; Institute for Social Research, University of Michigan, Ann Arbor, Michigan, USA; Knight Cancer Institute, Oregon Health & Science University, Portland, Oregon, USA; Federal University of Sao Paulo; Department of Molecular Biology, Princeton University, Princeton, NJ, USA; University of Rochester Medical Center, Departments of Psychiatry, Psychology, Neuroscience, and Obstetrics and Gynecology; Yale Child Study Center, Yale School of Medicine, New Haven CT, USA; Murdoch Childrens Research Institute, Royal Childrens Hospital, Flemington Road, Parkville, Victoria 3052, Australia; Department of Psychiatry, Universidade Federal do Rio Grande do Sul UFRGS, Porto Alegre, Brazil; Developmental Imaging, Murdoch Childrens Research Institute, Melbourne, Australia.; School of Psychology and Centre for Social and Early Emotional Development SEED, Deakin University, Geelong, Australia.; SAMRC Unit on Risk & Resilience in Mental Disorders, Department of Psychiatry and Neuroscience Institute, University of Cape Town, Cape Town, South Africa; Department of Paediatrics & Child Health, Red Cross Childrens Hospital and SA-MRC Unit on Child & Adolescent Health, University of Cape Town, South Africa

**Keywords:** Epigenetics, DNA methylation, MRI, early life, longitudinal, multi-cohort analysis

## Abstract

Epigenetic processes, such as DNA methylation, show potential as biological markers and mechanisms underlying gene-environment interplay in the prediction of mental health and other brain-based phenotypes. However, little is known about how peripheral epigenetic patterns relate to individual differences in the brain itself. An increasingly popular approach to address this is by combining epigenetic and neuroimaging data; yet, research in this area is almost entirely comprised of cross-sectional studies in adults. To bridge this gap, we established the Methylation, Imaging and NeuroDevelopment (MIND) Consortium, which aims to bring a developmental focus to the emerging field of Neuroimaging Epigenetics by (i) promoting collaborative, adequately powered developmental research via multi-cohort analyses; (ii) increasing scientific rigor through the establishment of shared pipelines and open science practices; and (iii) advancing our understanding of DNA methylation-brain dynamics at different developmental periods (from birth to emerging adulthood), by leveraging data from prospective, longitudinal pediatric studies. MIND currently integrates 15 cohorts worldwide, comprising (repeated) measures of DNA methylation in peripheral tissues (blood, buccal cells, and saliva) and neuroimaging by magnetic resonance imaging across up to five time points over a period of up to 21 years (N_pooled DNAm_ = 11,299; N_pooled neuroimaging_ = 10,133; N_pooled combined_ = 4,914). By triangulating associations across multiple developmental time points and study types, we hope to generate new insights into the dynamic relationships between peripheral DNA methylation and the brain, and how these ultimately relate to neurodevelopmental and psychiatric phenotypes.

## Background

DNA methylation (DNAm) is an epigenetic process that can regulate gene activity in response to both genetic and environmental influences [1, 2], beginning *in utero*. While DNAm plays a key role in *healthy* development and function [3], alterations in DNAm have also been linked to the emergence of *disease states*, including brain-based disorders such as neurodevelopmental, psychiatric, and neurological conditions [4–10]. In addition, peripheral markers of DNAm are often more accessible to measure than several brain-based phenotypes. Together, these properties make DNAm a promising molecular system in the search for both biological markers and mechanisms underlying gene-environment impacts on brain-based disorder and phenotypes. However, we still know little about how peripheral DNAm patterns relate to individual differences in the brain structure and function itself – the target organ of interest.

To address this gap, a growing number of researchers have examined associations between DNAm and brain features (typically measured using magnetic resonance imaging; MRI), giving rise to the new field of *Neuroimaging Epigenetics* [11–13]. Already more than 100 articles combining DNAm and brain MRI have been published to date, but important limitations remain. Most studies have assessed DNAm and the brain only at a single (cross-sectional) time point, are based on adult samples (∼80%), have small to moderate sample sizes (median *N* = 98, range 14-715), and have primarily used a candidate gene approach (i.e., focusing on a limited set of CpGs; 67%). The first large-scale multi-cohort investigations using an epigenome-wide approach are now emerging; these generally show modest associations, with for example only two CpGs in blood found to be associated with hippocampal volume in a pooled analysis of 3,337 participants after genome-wide correction [14].

## Why was the consortium set up?

Rapid advances in Neuroimaging Epigenetics have also been accompanied by several key challenges. First, studies in this field have been highly heterogeneous in terms of their design, characteristics, and methodology, with few shared practices, limiting comparability between findings and the identification of robust neuroimaging epigenetic associations. Second, the reliance on cross-sectional measurements, mainly in adults, likely ignores important temporal dynamics, as both DNAm and brain structure and volume are known vary over time [15–19] and in different ways (i) depending on the region examined (i.e., where in the genome or in the brain), (ii) in terms of their developmental trajectory (i.e., linear or non-linear), (iii) in light of (sensitive) periods with more change/growth (e.g., childhood and adolescence), and (iv) in association with different downstream phenotypes (e.g., neurodevelopmental, psychiatric). For example, recent evidence points to epigenetic *‘timing effects*’ [6–8], whereby DNAm patterns in cord blood at birth have been shown to prospectively associate with certain neurodevelopmental problems in childhood (e.g., ADHD symptoms [6, 7], social communication deficits [8]) *more strongly* than DNAm patterns measured cross-sectionally during childhood. Similar ‘timing effects’ have also been reported for changes in brain volume, with the strength of phenotypic associations varying depending on timing of assessment [20]. Yet, how DNAm and neuroimaging measures relate *to each other* at different developmental stages is largely unknown. Finally, studies to date are typically based on single cohorts with small sample sizes of *n*<100, with limited statistical power to detect what are likely to be small associations. While neighboring fields have already showed how large-scale collaborative efforts between cohorts [21–24] can be achieved, such initiatives at the intersection of epigenetics, imaging and development are lacking.

Recently, we established The Methylation, Imaging and NeuroDevelopment (MIND) Consortium to address these gaps. Our overarching goal is to understand how peripheral DNAm patterns relate to variation in brain structure and function across development. Specifically, we aim to bring a developmental focus to the emerging field of Neuroimaging Epigenetics by (i) promoting collaborative, adequately powered developmental research via multi-cohort analyses; (ii) increasing scientific rigor through the establishment of shared pipelines and open science practices; and (iii) elucidating directionality of associations between DNAm and the brain via the use of prospective, longitudinal cohorts across development (from birth through emerging adulthood).

## Who is in the consortium and what has been measured?

The MIND consortium brings together a global set of research teams and cohorts from North America, South America, Europe, African and Australia (**Figure 1**). It currently includes 15 cohorts, spanning population-based, twin and high-risk cohorts. The unifying feature of these cohorts is the availability of genome-wide DNAm data (Illumina 450K or Illumina EPIC chip) and neuroimaging data (e.g., T1-weighted images, diffusion-weighted images, and/or resting state functional MRI) collected at one or more time points across development, with at least one assessment prior to age 18 years. Most cohorts have data from fetal life into childhood, with several cohorts also covering adolescence and early adulthood. With ongoing data collection in many cohorts, new DNAm and neuroimaging data are also expected to become available at additional time points. Key characteristics of cohorts included in the MIND consortium can be found in **Table 1 and Figure 2**. Full cohort descriptions and study-specific acknowledgements can be found in the **Supplementary materials.**

**Figure 1.**
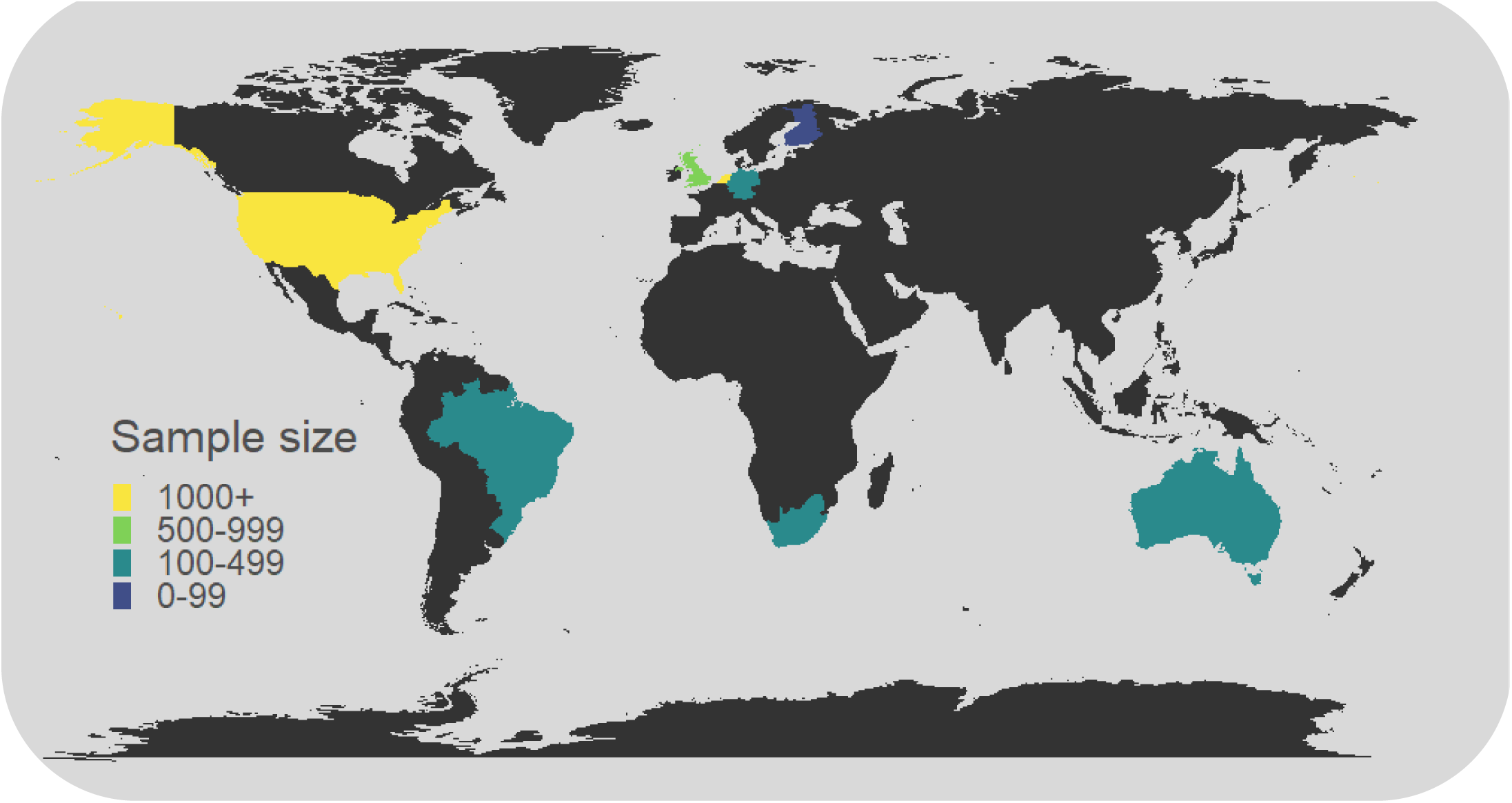
World map of sample size per country (overlap DNAm and neuroimaging), as covered by MIND. Sample sizes reflect *expected* sample sizes after sample processing (i.e., those featured in Table 1, but additionally including expected sample sizes for BHRC (*N* = 430), MTWinS (*N* = 708), and (*N* = 100)).

**Table 1.**
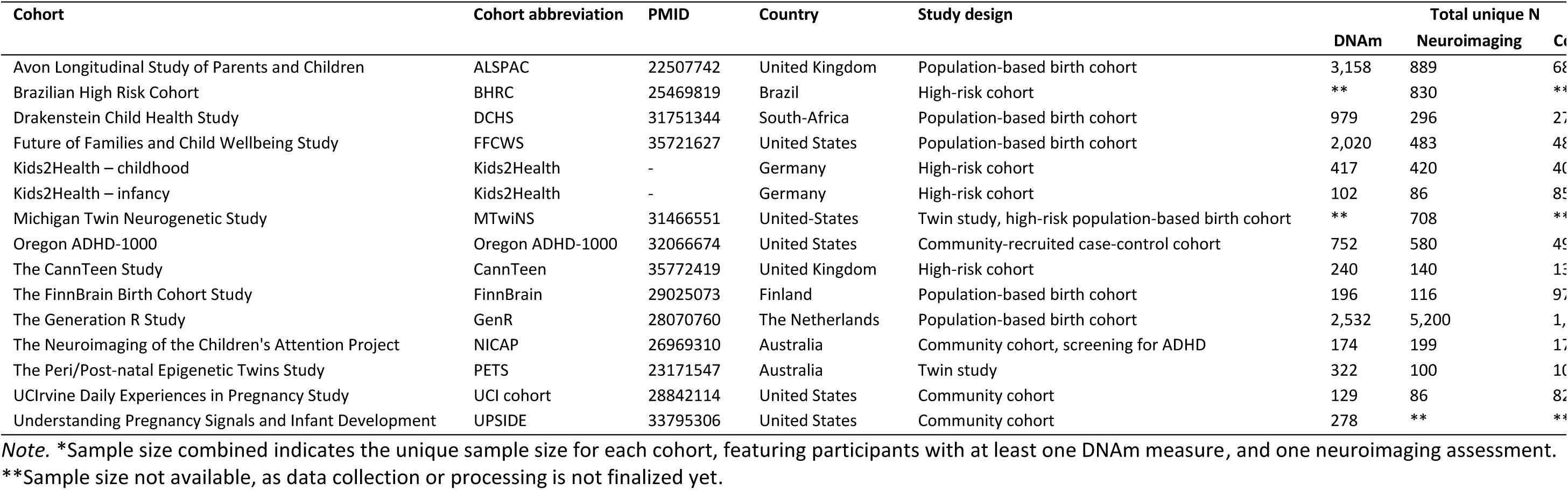
Key characteristics of cohorts participating in MIND

**Figure 2.**
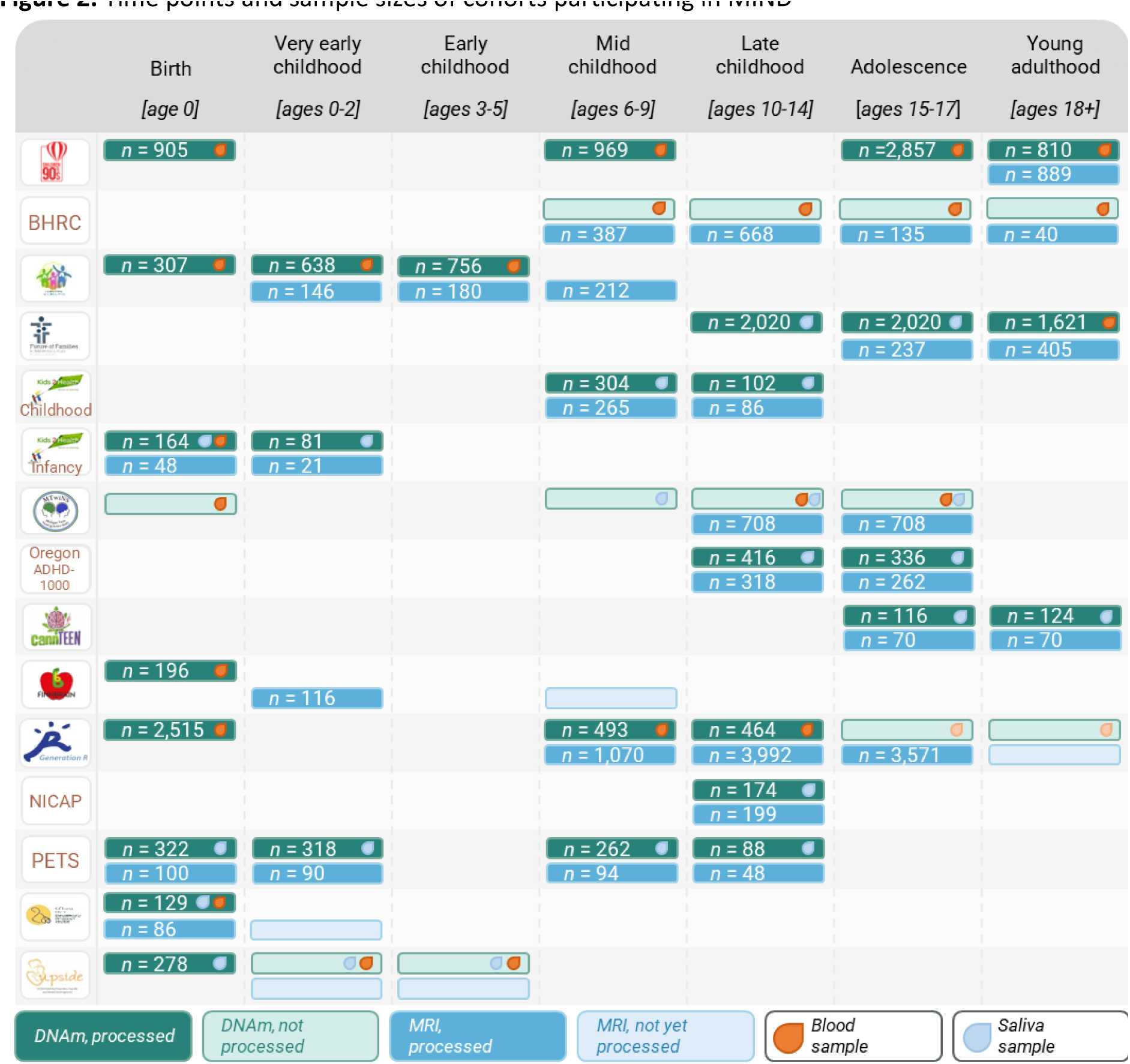
Time points and sample sizes of cohorts participating in MIND

To date, the MIND cohorts comprise a total of 11,299 participants with DNAm profiles, 10,133 participants with neuroimaging data, and 4,914 participants with both data types for neuroimaging epigenetic analyses. This is a significant expansion in sample size compared to the current average for *developmental* neuroimaging epigenetic studies (median *N* = 80, range 33-715 [11]). While a sample of 80 is only sufficiently powered (80%) to detect associations of medium effect (Cohen’s *d* = 0.45), with MIND we will be able to also detect associations of small effect (Cohen’s *d* = 0.08) [25], enabling us to characterize time-varying DNAm-brain associations in a more robust and nuanced manner. All cohorts have at least one time point of measurement, but 93% have two or more, up to five repeated time points. Cohorts participating in MIND are also notably diverse, including participants from various backgrounds in terms of ethnicity, geographical location and socio-economic environments, enabling the investigation of neuroimaging epigenetic associations across different settings, and increasing the inclusivity and generalizability of the findings. In addition, participants from most of the included cohorts have undergone extensive environmental, molecular and phenotypic profiling. As shown in **Table 2**, all cohorts have genetic data of participants – and in some cases also for relatives – which can be leveraged to account for population stratification, calculate polygenic scores, investigate gene-environment correlations and interactions as well as in some cases to study genetic nurture effects (i.e., using trio genetic data). Further, most cohorts have data on developmental and (mental) health outcomes, typically comprising measures of neurodevelopment, behavior, cognition and psychiatric symptoms, but also often feature other phenotypes such as anthropometrics and physical health assessments. Most cohorts have also measured environmental exposures, typically beginning *in utero* for the birth-cohorts (e.g., maternal smoking, diet, psychopathology and psychosocial stress during pregnancy) and during childhood/adolescence (e.g., early life stressors, parental influences, home environment and broader socio-economic factors). Finally, almost all cohorts have data on additional biological markers (e.g., inflammation) and profiling of other *omics* (e.g., microRNAs, metabolomics, proteomics and the gut microbiome).

**Table 2.**
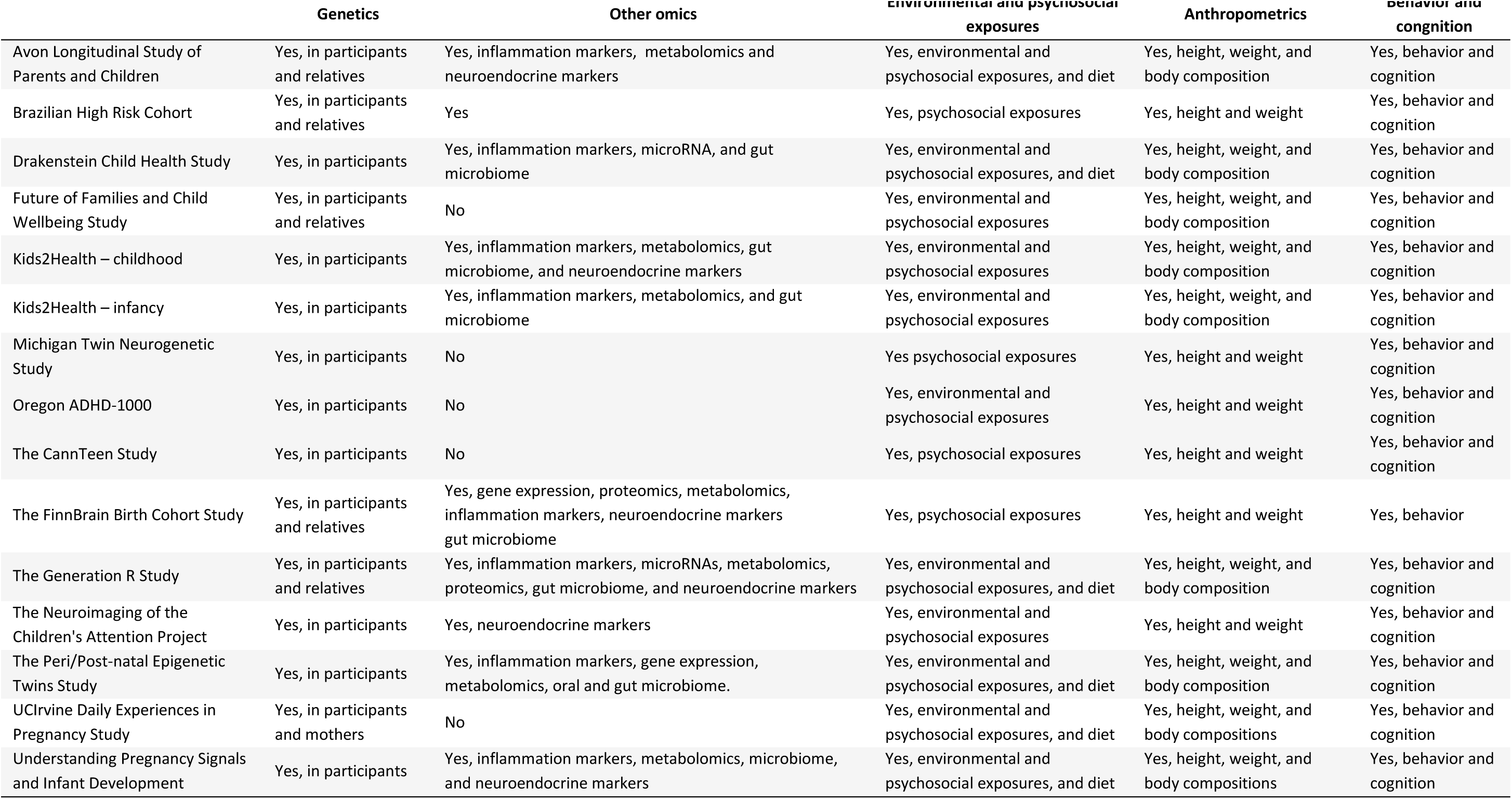
Overview of available measures different cohorts

## How do we work together?

MIND operates on a federated model, wherein methods, practices, and results are shared among consortium members, while maintaining the confidentiality of the underlying individual-level data. Contributors have the flexibility to propose new projects, which operate on an opt-in basis. Participation from different cohorts in each project is optional and contingent on data availability, data sharing policies, resource allocation, and cohort-specific research interests. Similarly to consortia such as the Pregnancy And Childhood Epigenetics (PACE) Consortium [22], project leads are responsible for producing detailed analysis plans, which wherever possible will be pre-registered for transparency, and project leads will also be responsible for developing and testing analysis scripts. In the case of meta-analytic projects, scripts are disseminated across the consortium by the project lead for implementation by participating cohorts, who then share cohort-specific summary statistics with the project lead for pooling of results. Options for mega-analyses (i.e., pooled analyses of individual participant data) will also be explored in light of the increasing availability and uptake of federated data sharing platforms.

## Getting started: Identifying and addressing challenges in collaborative research on developmental neuroimaging epigenetics

Modeling (potentially) time-varying associations between two high-dimensional data modalities within a consortium framework offers new opportunities but also considerable challenges. Here, we outline three anticipated challenges within MIND, and reflect on potential strategies that could be used to address them (Figure 3).

**Figure 3.**
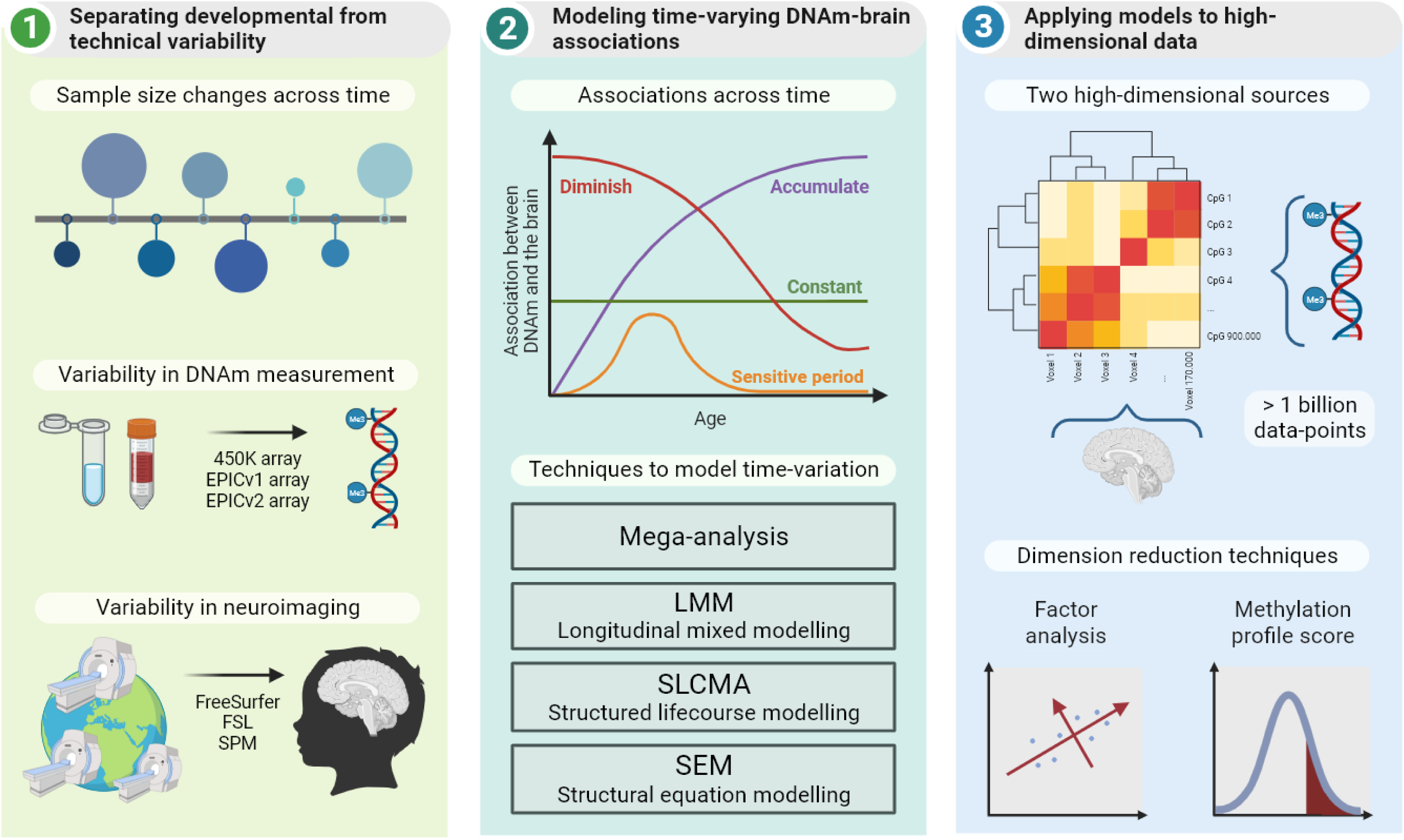
Key challenges in Neuroimaging Epigenetics

### Challenge #1: Separating developmental from technical variability

A key goal of MIND is to characterize neuroimaging epigenetic associations across development. We therefore plan to integrate data from cohorts covering different ages and developmental periods. While all cohorts in MIND have collected DNAm and neuroimaging data using comparable techniques, they still show substantial variability in sample characteristics and data processing methods. Notably, technical variability could increase generalizability of findings, e.g., MIND findings are more generalizable when using a range of methods, whereas studies using a single technique alone may be less generalizable to cases where different techniques are used. Yet, technical variability also complicates integration of different findings across cohorts. Furthermore, sources of technical variability across cohorts are tied to (or coincide with) differences in the timing of assessments, making it difficult to separate (developmental) signal from (technical) noise. This raises the challenge of balancing the need of maximizing statistical power (typically by increasing pooled sample size as much as possible) with maximizing comparability across diverse cohorts (instead calling for more focused pooling of data based on shared characteristics). We highlight below main sources of variability in sample characteristics, DNAm, and neuroimaging.

#### Sample characteristics

MIND cohorts vary widely with respect to *sample size*, from numerous smaller studies with neonatal MRI (e.g., *N* = 86, UCI cohort) to larger cohorts with MRI data at different time points in childhood and adolescence (*N* = 822 at around age 18, ALSPAC cohort). This variation in sample size leads to uneven data availability across different developmental periods, resulting in unbalanced representation and differences in statistical power between these developmental periods. Furthermore, while almost all cohorts have repeated measures of DNAm or neuroimaging (or both), a small number have only a single time point available. This discrepancy results in *partial overlap* of samples when analyzing different developmental periods. In other words, the same group of participants is not consistently tracked across all time points in every study. To complicate matters, the ‘spacing’ of time points between DNAm and neuroimaging differs between cohorts. For example, while some cohorts have both DNAm and neuroimaging available in the neonatal period (maximum gap of a few weeks), others have DNAm at birth but only started with neuroimaging assessments in mid-childhood, resulting in a gap of several years. To address these sample differences, careful consideration of covariates (e.g., age at DNAm and neuroimaging assessments and the time gap between them) as well as methods to address partial sample overlap and sample size imbalance across cohorts (e.g., through the use of weighted approaches and leave-one-out analyses to test the stability of results) will be needed.

#### DNAm

Another source of variability across studies is in how DNAm is measured, both in terms of DNAm array and tissue examined. The type of DNAm array used varies not only *between* MIND cohorts but at times also *within* longitudinal cohorts. The gold standard for DNAm assessment is whole-genome bisulfite sequencing (WGBS), offering single-nucleotide precision and covering about 95% of all CpGs on the genome, or roughly 28 million CpG sites [26]. However, its high cost and computational burden limits its use in large-scale cohorts. As an alternative, Illumina has developed a series of more affordable BeadChip microarrays, starting with the HumanMethylation27 BeadChip in 2008, expanding to over 450,000 CpGs with the 450K array in 2011 [27], and further to over 850,000 CpGs with the EPICv1 in 2016 [28]. The latest version, EPICv2, features an additional 186,000 CpGs [29]. The cost-effective nature of these arrays (∼3 times cheaper than WGBS) has made it possible for cohorts to measure hundreds of thousands of CpGs across the genome in larger samples, and as a result these methods have been eagerly adopted by many cohort studies. Yet, DNAm arrays and their technology change over time, and while these changes are marked by the removal of poor-quality sites [30] and addition of new probes with evidence for biological or clinical importance, the availability of data across several arrays presents challenges for longitudinal cohorts transitioning between versions. The discrepancies at the individual CpG level could reduce consistency in longitudinal research [31, 32]. Consequently, it may be difficult for longitudinal cohorts to distinguish between developmental changes *vs* variations due to array or batch-related factors, especially as measuring the same sample using multiple arrays is hardly ever done. Fortunately, studies comparing arrays have reported generally high concordance [29, 31–33].

DNAm patterns show limited comparability across tissues [34–36]. This poses a challenge in developmental epigenetic research, where - for example - samples at birth are often obtained from cord blood (or other accessible tissues such as placenta), as compared to peripheral blood, saliva, or buccal tissue at later time points. In addition, cohorts typically collect DNAm in only one tissue type, although MIND does feature a small subset of cohorts that collected DNAm in multiple tissues (e.g., saliva and blood, Kids2health). A common practice for many multi-cohort studies (e.g., [6, 14, 37]), has been to focus on blood tissue, including cord, heel prick and peripheral whole blood during development. While utilizing data obtained from a single tissue can enhance comparability and reduce noise between studies, it can come at the cost of lower sample size and limited generalizability to different tissues. Yet, even within the same tissue, cell-type composition can change dramatically across development, both in terms of the type and proportion of cells in bulk tissue. For example, nucleated red blood cells (nRBCs) are abundant in cord blood, but largely absent in blood later in development. Cell-type composition in bulk tissue can be adjusted for using age/tissue-appropriate reference panels (e.g., reference for cord blood versus peripheral blood at later time points, [6, 37]). However, this also introduces a further source of technical variability in longitudinal analyses (given that each panel has been estimated using different data sources). For example, a previous study characterizing DNAm trajectories across development found widespread age-related changes in DNAm patterns over the first two decades of life [15]. In some cases, these changes were non-linear (e.g. DNAm changing more rapidly from birth to mid-childhood, compared to later time points), raising questions about whether these changes are due to developmental timing or differences between cord and peripheral blood. In this respect, the creation of a single, integrated panel which estimates a more comprehensive set of cell-types, including (i) cells that may be present at only one time point (e.g., nRBCs at birth), as well as (ii) cell types that may be ubiquitous across development, but still vary in abundance over time (e.g., naïve versus memory B cells), could offer an attractive solution to better separate tissue from timing-related differences.

#### Neuroimaging

Measurement heterogeneity is also a factor for neuroimaging assessments. MRI scanner types typically vary across study sites, and even within *one* site, scanners are often upgraded or replaced over time. Different strategies that have been proposed for reducing this unwanted geographical and temporal variation, include harmonization techniques like ComBat [38–40], deep learning [41], or hierarchical Bayesian regression [42]. However, removing the geographical and temporal variation from neuroimaging assessments can be challenging in developmental research: consider participants scanned repeatedly at different ages (e.g., as newborns and later in development) at various locations or with different scanners. While harmonization techniques aim to eliminate unwanted scanner or site variations, there is a risk they might also inadvertently remove developmental variation-of-interest. Currently, there is a lack of consensus about whether these methods should or should not be used for developmental cohorts. For example, one large-scale study by Ge et al. [19], involving almost 40,000 participants from 86 cohorts, used ComBat to control for unwanted site effects while comparing statistical methods for normative modeling of brain morphometric data. Another large-scale study by Kia et al., [42], also including almost 40,000 participants from 79 scanners, harmonized their data by using federated hierarchical Bayesian regression. In contrast, another large study by Bethlehem et al. [18], including over 100,000 participants from 100 cohorts, chose not to perform harmonization techniques for constructing normalized brain charts. Overall, it is unclear how best to strike a balance between reducing unwanted scanner variation while also preserving developmental changes crucial for accurate interpretation.

Most neuroimaging processing software packages - such as FreeSurfer, Statistical Parametric Mapping (SPM), and FMRIB Software Library (FSL) – have been developed for and tested in *adult* samples, raising concerns about how these methods perform in data from *infants and children*. For example, although the standard FreeSurfer suite has been successfully applied to data from children aged 4 years [43], *Infant FreeSurfer* has been developed for infants aged 0-2 [44]. Other software packages developed especially for fetal or neonatal brain scans include iBEAT [45] and NEOCIVET [46]. An important question that follows is whether cohorts should prioritize methodological consistency (i.e., reduce technical variability at the cost of potentially lower accuracy, by processing all neuroimaging data with the same software) or developmental sensitivity (i.e., use accurate tools at the cost of methodological consistency, by processing neuroimaging data with developmentally appropriate software) when harmonizing cross-cohort neuroimaging data across different life stages. Both strategies have been used in the past. For example, a recent analysis from the ENIGMA Lifespan consortium, including data from age 3 to 90 years across 86 cohorts, only used standard FreeSurfer [19]. In contrast, Bethlehem et al. [18] created normalized brain charts for mid-gestation up to age 100, while allowing for the use of developmentally-sensitive processing software across cohorts. Consequently, all adult cohorts were processed using FreeSurfer, but there was considerable heterogeneity of methods in data from pre-birth to early childhood (e.g., manual segmentation, NEOCIVET, customized FreeSurfer versions, Infant FreeSurfer, and standard Freesurfer). It is still unclear whether the use of age-tailored software tools adds or reduces noise in longitudinal analyses that span several developmental periods (e.g., birth to early adulthood). Other strategies include leveraging age-appropriate software, while clustering analyses by developmental period, and accounting for software-related variations during the meta-analysis phase. Alternatively, findings can be triangulated using different processing software. Overall, research evaluating the best options for leveraging developmentally sensitive data is needed to provide evidence-based guidelines for the field.

### Challenge #2: Modeling time-varying DNAm-brain associations in multi-cohort analyses

Besides considering how best to account for variability between MIND cohorts, decisions must also be made regarding how best to statistically perform multi-cohort integration. In consortium settings, standard meta-analyses remain the most popular option, where an exposure (e.g., DNAm at specific loci) is associated with an outcome (e.g., a brain feature) using regression models within each individual cohort, after which cohort-level summary statistics are pooled through meta-analysis. Such methods were used by Jia et al. [14], which is currently the largest DNAm-brain multi-cohort study [12], meta-analyzing data from 3,337 participants (mainly adults) across 11 cohorts. Notably, only two CpGs were found to be associated with hippocampal volume after correction for multiple testing, and none for the thalamus or the nucleus accumbens – the other two regions investigated. While this could suggest that the meta-analysis was either underpowered to detect modest effects or that peripheral DNAm and volumetric brain differences are largely unrelated, it could also indicate that rather than being constant, associations vary over time. Unless explicitly modeled, however, this information is lost in standard meta-analytic approaches, which inherently assume constant exposure-outcome associations; i.e., that the link between DNAm and the brain does not change over time. Although this restriction could have the advantage of identifying age-averaged associations (which may be more generalizable across age groups), it comes at the cost of obscuring time-varying effects. This may be particularly problematic for studies which pool data from both pediatric and adult cohorts, as changes in DNAm patterns and brain features are expected to be more drastic earlier in life.

An alternative approach to address developmental timing in multi-cohort studies involves strategic pooling of summary data at specific age ranges, as opposed to pooling all cohorts together regardless of age. This approach is commonly used in the PACE consortium (e.g., [6, 37]), prioritize one epigenetic time point of interest (e.g., birth, because it is generally the time point with the largest sample size, or because of a focus on pregnancy exposures) and then query later DNAm assessments by (i) adopting a follow-forward approach, where significant CpG sites are selected for further analysis and tested at subsequent time points (e.g., [7, 9, 47]) or by (ii) conducting a completely new set of analyses at later developmental stages (e.g., running separate epigenome-wide analyses using DNAm at birth and DNAm in childhood in relation to the same child outcome [6, 37]). While this strategy helps to partition analyses according to more developmentally-specific age groups, it does not directly quantify temporal changes in DNAm-brain associations, nor does it account for the potential non-linearity of this relationship over time. For this, meta-regression methods could be applied, which allow to test, within the same model (still based on cohort-specific summary statistics), (i) how DNAm and brain features associate at each available time points/developmental period, (ii) whether these associations change across time periods, and (iii) whether changes are linear or non-linear [15–17]. For an example of applying longitudinal meta-regression to quantify DNAm timing effects on child health outcomes using cohort-level summary statistics from the PACE Consortium, see [48].

While the above strategies can help us get started, ultimately, the transition toward a *mega*-analysis framework using individual-level data will offer greater modeling flexibility and new opportunities to address developmental questions in collaborative multi-cohort studies. Three concrete examples of potential applications here include longitudinal mixed models (LMM), structured life-course modeling analysis (SLCMA) and structural equation modeling (SEM). Mixed models test whether *changes* in a predictor (measured at repeated time points, e.g., DNAm at birth, age 6, and age 10) associates with an outcome (measured at single time points, e.g., brain feature at age 14) over time. While we are not aware of any existing *multi*-cohort studies using LMM to model DNAm-brain associations, this method was recently applied (i) in a single-cohort study to examine whether DNAm sites across time prospectively associate with the amygdala:hippocampus ratio in early adulthood [49], and (ii) to individual-level data from two longitudinal cohorts in order to characterize how epigenome-wide DNAm patterns vary over the first two decades of life [15]. As an alternative to LMM, SLCMA uses repeated measures of a *predictor* to test competing temporal hypotheses about its effect on a particular outcome (measured at a single time point). It has been used for example to test the developmental effect of stress exposure on child DNAm, showing that stressful experiences are more likely to associate with changes in DNAm if they occur early rather than late in childhood [50, 51]). This approach could be extended to model repeated DNAm as a predictor, in order to establish whether epigenetic effects on brain outcomes may also be developmentally-specific (orange line, Figure 3), or might diminish or accumulate over time (red and purple lines, Figure 3), as opposed to being temporally invariant (green line, Figure 3). Finally, SEM allows modeling of repeated measures for *both* the predictor and the outcome; it has been previously applied to DNAm and neuroimaging data, although not within the same study. In the context of MIND, this model could be used to integrate DNAm-brain data, for example, to (i) estimate the degree of temporal stability in either epigenetic patterns and brain features across development (i.e., autoregressive paths), (ii) examine relationships between these DNAm and the brain over time (i.e., cross-lagged paths), and (iii) to test genetic/environmental predictors or phenotypic (health) outcomes of identified DNAm-brain associations, as well as mediating effects (i.e., indirect paths). Despite the potential of these advanced methods to model complex (repeated) DNAm-brain measures for time-varying associations, their implementation within a multi-cohort framework remains a more distant goal. Success in this respect will first depend on overcoming existing barriers in sharing of individual-level data, increasing availability of meta-analyses for advanced statistical models, accessing a large enough set of cohorts with sufficiently comparable repeated DNAm/neuroimaging measures, and deciding how best to balance the complexity of longitudinal methods with the high-dimensionality of these data types, as discussed in more detail below.

### Challenge #3: Analyzing high-dimensional data

A third set of challenges relates to the dimensionality of DNAm and MRI data. DNAm data is commonly investigated through univariate epigenome-wide associations (with covariate adjustment), where DNAm-trait relationships are studied for a large number of individual sites across the genome. Epigenome-wide investigations currently interrogate up to 900,000 data points per person, depending on which array is used (note that even more data points can be measured with some technologies [26]). Brain data are often examined voxel-wide, where local density of gray matter is compared between participants (e.g., per 1mm_3_ of the brain), again resulting in hundreds of thousands data points per participant. As such, if epigenome- and voxel-wide data are analyzed together in the same model, even if just at a single time point, this would add up to examining more than a billion associations (CpG by voxel). Such models, especially if extended longitudinally, are currently not feasible (due to limited power) or practical (due to computational burden), therefore requiring some level of data manipulation.

One common approach to handling high-dimensional data is to apply dimension reduction techniques, such as principal component analysis, (parallel) independent component analysis, local Fisher’s discriminant analysis, and canonical correlation analysis [52, 53]. These methods have indeed been applied to neuroimaging epigenetic data in single cohorts, although their application to multi-cohort analyses can be challenging due to the potential of obtaining different model solutions (e.g., factors or components comprised of different CpGs/brain features, or different weights) in different cohorts. Alternatively, we can condense high-dimensional data into singular scores that can be computed in a comparable way across cohorts, as done for example in the case of biological age estimates such as epigenetic age and brain age [54, 55]. In particular, the use of methylation profile scores (MPS) is gaining popularity in epigenetic research [56], following in the footsteps of polygenic scores (PGS) within the field of genetics. Within MIND, we could use MPSs to proxy at an epigenetic level brain-relevant exposures (e.g., prenatal smoking or insufficient sleep), biological processes (e.g., inflammation, neuroendocrine function), and health outcomes (e.g., mental or cardiometabolic phenotypes). We could also seek to develop MPSs of brain features themselves, an approach not yet attempted. Another application of MPSs is to use them as a proxy for missing covariates or to enhance the quality of data imputation, which could be particularly helpful in the case of multi-cohort analyses (as certain cohorts may not have collected important covariates, or at least not at the specific age of interest). Overall, MPSs may offer a practical solution to manage high dimensionality while also aggregating together many epigenetic loci of small effect; yet, they also have their challenges. For example, MPSs are increasingly constructed using penalized linear regression techniques, which require adequate consideration and availability of training, tuning and testing datasets. The selection and splitting of datasets, and the amount of data reserved for training versus testing, can reduce sample size and power. Of note, to obtain reliable estimates for PGSs in complex traits, discovery samples from tens to hundreds of thousands of individuals have been required. Sample sizes required to construct reliable MPSs remain undefined. Furthermore, although the use of aggregate scores can help increase the amount of explained variance, it comes at the cost of not being able to identify specific loci, genes or biological pathways, needed for identifying causal pathways for biomedical interventions. Finally, the predictive power of PRSs is influenced by ancestry [57]. While some evidence shows this may be less of an issue with MPSs [58], it will be of interest to test performance of MPSs in MIND to generate MPSs that can ideally be applied across ancestries.

## Conclusions

The potential of large-scale collaborative efforts between cohorts has been clearly demonstrated by the success of genome-wide association studies (for example by the Psychiatric Genetic Consortium [21]), epigenome-wide association studies (for example by PACE [22]), and neuroimaging studies (for example by ENIGMA [23] or specifically ENIGMA-ORIGINs [24]). The MIND consortium aims to apply these principles to the emerging field of neuroimaging epigenetics, with a particular emphasis on development. Priorities include fostering multi-cohort analyses to access collaborative, adequately powered developmental research; to establish shared pipelines; and to elucidate the time-varying relationship between peripheral DNAm and brain development through prospective, longitudinal cohorts spanning pre-birth to young adulthood. MIND is committed to pursuing these goals while supporting open science practices, including the use of pre-registration of studies, sharing of analysis scripts, and making full results (e.g., meta-analysis summary statistics) openly accessible. Through these activities, the consortium aims to facilitate an integrative and efficient research environment and to enhance the transparency and reproducibility of our research. Overall, we hope that MIND will represent a significant step towards shedding light into the complex, dynamic relationship between peripheral DNAm and the brain, and to understanding how these ultimately relate to neurodevelopmental and psychiatric phenotypes.

## Joining us

The MIND Consortium operates as an open network, welcoming researchers interested in joining with one or more of its cohorts. In this consortium, each participant manages and analyses their data locally. Detailed protocols for each study can be accessed through their individual websites (see **Supplementary materials**) or by contacting the study investigators. In order to participate in MIND, cohorts must feature at least one assessment of DNAm (peripheral, array-based) and neuroimaging collected before the age of 18 years. Those interested in collaborating with the MIND Consortium are encouraged to contact the corresponding authors. More information can also be found on our website: https://www.erasmusmc.nl/en/research/groups/methylation-imaging-and-neurodevelopment-mind-consortium#

## Funding and acknowledgments

The work of CAMC is supported by the European Union’s HorizonEurope Research and Innovation Programme (FAMILY, grant agreement No 101057529; HappyMums, grant agreement No 101057390) and the European Research Council (TEMPO; grant agreement No 101039672). This research was conducted while CAMC was a Hevolution/AFAR New Investigator Awardee in Aging Biology and Geroscience Research. EW and CAMC are supported by the European Union’s Horizon 2020 Research and Innovation Programme (EarlyCause, grant agreement No 848158). EW, MS and VB received funding from UK Research and Innovation (UKRI) under the UK government’s Horizon Europe / ERC Frontier Research Guarantee [BrainHealth, grant number EP/Y015037/1]. EW also received funding from the National Institute of Mental Health of the National Institutes of Health (award number R01MH113930).

We are extremely grateful to all the families who took part in this study, the midwives for their help in recruiting them, and the whole ALSPAC team, which includes interviewers, computer and laboratory technicians, clerical workers, research scientists, volunteers, managers, receptionists and nurses. The UK Medical Research Council and Wellcome (Grant ref: 217065/Z/19/Z) and the University of Bristol provide core support for ALSPAC. GWAS data was generated by Sample Logistics and Genotyping Facilities at Wellcome Sanger Institute and LabCorp (Laboratory Corporation of America) using support from 23andMe. This publication is the work of the authors and they will serve as guarantors for the contents of this paper. A comprehensive list of grants funding is available on the ALSPAC website (http://www.bristol.ac.uk/alspac/external/documents/grant-acknowledgements.pdf). The Brazilian High-Risk Cohort – Happy Mums is supported by the National Institute of Developmental Psychiatry for Children and Adolescents, a science and technology institute funded by the Conselho Nacional de Desenvolvimento Científico e Tecnológico (CNPq; National Council for Scientific and Technological Development; grant number 573974/2008-0), Fundação de Amparo à Pesquisa do Estado de São Paulo (FAPESP; Research Support Foundation of the State of São Paulo; grant number 2008/57896-8), and Horizon – Health, through the project Understanding, predicting, and treating depression in pregnancy to improve mothers and offspring mental health outcomes (grant number 101057390). The Drakenstein Child Health study is funded by the Bill and Melinda Gates Foundation [grant number OPP 1017641], National Institute on Alcohol Abuse and Alcoholism [grant numbers R21AA023887, R01AA026834-01], US Brain and Behavior Research Foundation [grant number 24467], Collaborative Initiative on Fetal Alcohol Spectrum Disorders (CIFASD) [grant number U24 AA014811], South African Medical Research Council, UK Government’s Newton Fund [grant number NAF002/1001], Wellcome Trust [grant number 203525/Z/16/Z], South Africa’s National Research Foundation [grant numbers 105865, 120432], ABMRF/The Foundation for Alcohol Research, and Harry Crossley Foundation. FFCW data collection is funded by National Institutes of Health (NIH) R01-HD-036916 and a consortium of other funders. Methylation analyses are funded by the NIH: R01 HD076592, R01 MH103761, R01HL149869, and R01 MD011716. Brain imaging (SAND) is funded by the NIH: R01 MH103761 and R01 MH121079. Kids2health is funded by BMBF 01GL1743A. MTwiNS was supported by funds from the National Institute of Mental Health of the National Institutes of Health (NIH): UH3MH114249 & R01 MD081813; the Eunice Kennedy Shriver National Institute of Child Health and Human Development of the NIH: R01HD093334 & R01 HD104297 & R01 HD066040; the Brain and Behavior Foundation: NARSAD young Investigator Grant, The Avielle Foundation, and Institutional funds from the University of Michigan and Michigan State University. Any opinions, findings, and conclusions or recommendations expressed in this material are those of the authors and do not necessarily reflect the views of the NIH. The authors would like to thank the staff of the Twin Study of Behavioral and Emotional Development— Child (TBED-C) and Michigan Twins Neurogenetics Study (MTwiNS) studies for their hard work, and the authors thank the families who participated in TBED-C and MTwiNS for sharing their lives with us. The Oregon ADHD-1000 cohort is supported by grants from the National Institute of Mental Health of the National Institutes of Health under award numbers R37MH059105 (JTN), R01MH099064 (JTN), R01MH115357 (JTN), R01MH131685 (JTN, MAM). The CannTeen study was funded by the UK Medical Research Council (MR/P012728/1). The FinnBrain study was supported by the Academy of Finland, the Sigrid Juselius Foundation, the Signe and Ane Gyllenberg Foundation, the Jane and Aatos Erkko, and the Foundation and State Grants for Clinical Research (ERVA). The general design of the Generation R Study is made possible by financial support from the Erasmus MC, Erasmus University Rotterdam, the Netherlands Organization for Health Research and Development and the Ministry of Health, Welfare and Sport. The EWAS data were funded by a grant from the Netherlands Genomics Initiative (NGI)/Netherlands Organisation for Scientific Research (NWO) Netherlands Consortium for Healthy Aging (NCHA; project nr. 050-060-810), by funds from the Genetic Laboratory of the Department of Internal Medicine, Erasmus MC, and by a grant from the National Institute of Child and Human Development (R01HD068437). We gratefully acknowledge the contribution of children and parents, general practitioners, hospitals, midwives and pharmacies in Rotterdam. The NICAP cohort is supported by the National Health and Medical Research Council of Australia (NHMRC #1065895 and #2029361) and a grant from the Waterloo Foundation. PETS was supported by grants from the Australian National Health and Medical Research Council (#437015, #607358, #114333), the Bonnie Babes Foundation (#BBF20704), the Financial Markets Foundation for Children (#032-2007), and the Victorian Government’s Operational Infrastructure Support Program. The UCIrvine Daily Experiences in Pregnancy Study was funded in part by a European Research Area Network (ERA Net) Neuron grant MecTranGen 01EW1407A to C.B. and National Institutes of Health grants R01 HD-060628 and R01 MD-017387.

## Data Availability

All data produced in the present study are available upon reasonable request to the authors

## References

1. Szyf, M., P. McGowan, and M.J. Meaney, The social environment and the epigenome. Environmental and molecular mutagenesis, 2008. 49(1): p. 46–60.

2. Van Dongen, J., et al., Genetic and environmental influences interact with age and sex in shaping the human methylome. Nature communications, 2016. 7(1): p. 11115.

3. Jones, M.J., S.J. Goodman, and M.S. Kobor, DNA methylation and healthy human aging. Aging cell, 2015. 14(6): p. 924–932.

4. Gao, X., et al., Epigenetics in Alzheimer’s disease. Frontiers in Aging Neuroscience, 2022. 14: p. 911635.

5. Abdolmaleky, H.M., et al., Methylomics in psychiatry: modulation of gene–environment interactions may be through DNA methylation. American Journal of Medical Genetics Part B: Neuropsychiatric Genetics, 2004. 127(1): p. 51–59.

6. Neumann, A., et al., Association between DNA methylation and ADHD symptoms from birth to school age: a prospective meta-analysis. Translational Psychiatry, 2020. 10(1): p. 398.

7. Walton, E., et al., Epigenetic profiling of ADHD symptoms trajectories: a prospective, methylome-wide study. Molecular psychiatry, 2017. 22(2): p. 250–256.

8. Rijlaarsdam, J., et al., Epigenetic profiling of social communication trajectories and co-occurring mental health problems: a prospective, methylome-wide association study. Development and psychopathology, 2022. 34(3): p. 854–863.

9. Luo, M., et al., DNA methylation at birth and lateral ventricular volume in childhood: a neuroimaging epigenetics study. Journal of Child Psychology and Psychiatry, 2023.

10. Caramaschi, D., et al., Epigenome-wide association study of seizures in childhood and adolescence. Clinical epigenetics, 2020. 12: p. 1–13.

11. Walton, E., et al., A systematic review of neuroimaging epigenetic research: calling for an increased focus on development. Molecular Psychiatry, 2023: p. 1–9.

12. Wheater, E.N., et al., DNA methylation and brain structure and function across the life course: A systematic review. Neuroscience & Biobehavioral Reviews, 2020. 113: p. 133–156.

13. Berger, A., How does it work?: Magnetic resonance imaging. BMJ: British Medical Journal, 2002. 324(7328): p. 35.

14. Jia, T., et al., Epigenome-wide meta-analysis of blood DNA methylation and its association with subcortical volumes: findings from the ENIGMA Epigenetics Working Group. Molecular psychiatry, 2021. 26(8): p. 3884–3895.

15. Mulder, R.H., et al., Epigenome-wide change and variation in DNA methylation in childhood: trajectories from birth to late adolescence. Human molecular genetics, 2021. 30(1): p. 119–134.

16. Oh, E.S. and A. Petronis, Origins of human disease: the chrono-epigenetic perspective. Nature Reviews Genetics, 2021. 22(8): p. 533–546.

17. Shaw, P., et al., Neurodevelopmental trajectories of the human cerebral cortex. Journal of neuroscience, 2008. 28(14): p. 3586–3594.

18. Bethlehem, R.A.I., et al., Brain charts for the human lifespan. Nature, 2022. 604(7906): p. 525–533.

19. Ge, R., et al., Normative Modeling of Brain Morphometry Across the Lifespan using CentileBrain: Algorithm Benchmarking and Model Optimization. bioRxiv, 2023: p. 2023.01. 30.523509.

20. Fuhrmann, D., L.J. Knoll, and S.-J. Blakemore, Adolescence as a sensitive period of brain development. Trends in cognitive sciences, 2015. 19(10): p. 558–566.

21. Sullivan, P.F., The psychiatric GWAS consortium: big science comes to psychiatry. Neuron, 2010. 68(2): p. 182–186.

22. Felix, J.F., et al., Cohort profile: pregnancy and childhood epigenetics (PACE) consortium. International journal of epidemiology, 2018. 47(1): p. 22–23u.

23. Thompson, P.M., et al., The ENIGMA Consortium: large-scale collaborative analyses of neuroimaging and genetic data. Brain imaging and behavior, 2014. 8: p. 153–182.

24. Alex, A.M., et al., Genetic influences on the developing young brain and risk for neuropsychiatric disorders. Biological psychiatry, 2023. 93(10): p. 905–920.

25. Grin, J.W., Calculating statistical power for meta-analysis using metapower.

26. Kurdyukov, S. and M. Bullock, DNA methylation analysis: choosing the right method. Biology, 2016. 5(1): p. 3.

27. Dedeurwaerder, S., et al., Evaluation of the Infinium Methylation 450K technology. Epigenomics, 2011. 3(6): p. 771–784.

28. Pidsley, R., et al., Critical evaluation of the Illumina MethylationEPIC BeadChip microarray for whole-genome DNA methylation profiling. Genome biology, 2016. 17(1): p. 1–17.

29. Noguera-Castells, A., et al., Validation of the new EPIC DNA methylation microarray (900K EPIC v2) for high-throughput profiling of the human DNA methylome. Epigenetics, 2023. 18(1): p. 2185742.

30. Sugden, K., et al., Patterns of reliability: assessing the reproducibility and integrity of DNA methylation measurement. Patterns, 2020. 1(2).

31. Olstad, E.W., et al., Low reliability of DNA methylation across Illumina Infinium platforms in cord blood: implications for replication studies and meta-analyses of prenatal exposures. Clinical Epigenetics, 2022. 14(1): p. 80.

32. Solomon, O., et al., Comparison of DNA methylation measured by Illumina 450K and EPIC BeadChips in blood of newborns and 14-year-old children. Epigenetics, 2018. 13(6): p. 655–664.

33. Fernandez-Jimenez, N., et al., Comparison of Illumina 450K and EPIC arrays in placental DNA methylation. Epigenetics, 2019. 14(12): p. 1177–1182.

34. Gunasekara, C.J., et al., A genomic atlas of systemic interindividual epigenetic variation in humans. Genome Biology, 2019. 20: p. 1–12.

35. Walton, E., et al., Correspondence of DNA methylation between blood and brain tissue and its application to schizophrenia research. Schizophrenia bulletin, 2016. 42(2): p. 406–414.

36. Edgar, R.D., et al., BECon: a tool for interpreting DNA methylation findings from blood in the context of brain. Translational psychiatry, 2017. 7(8): p. e1187–e1187.

37. Rijlaarsdam, J., et al., DNA methylation and general psychopathology in childhood: an epigenome-wide meta-analysis from the PACE consortium. Molecular Psychiatry, 2023. 28(3): p. 1128–1136.

38. Beer, J.C., et al., Longitudinal ComBat: A method for harmonizing longitudinal multi-scanner imaging data. Neuroimage, 2020. 220: p. 117129.

39. Fortin, J.-P., et al., Harmonization of cortical thickness measurements across scanners and sites. Neuroimage, 2018. 167: p. 104–120.

40. Radua, J., et al., Increased power by harmonizing structural MRI site differences with the ComBat batch adjustment method in ENIGMA. Neuroimage, 2020. 218: p. 116956.

41. Cetin-Karayumak, S., et al., Harmonized diffusion MRI data and white matter measures from the Adolescent Brain Cognitive Development Study. Scientific Data, 2024. 11(1): p. 249.

42. Kia, S.M., et al., Closing the life-cycle of normative modeling using federated hierarchical Bayesian regression. Plos one, 2022. 17(12): p. e0278776.

43. Ghosh, S.S., et al., Evaluating the validity of volume-based and surface-based brain image registration for developmental cognitive neuroscience studies in children 4 to 11 years of age. Neuroimage, 2010. 53(1): p. 85–93.

44. Zöllei, L., et al., Infant FreeSurfer: An automated segmentation and surface extraction pipeline for T1-weighted neuroimaging data of infants 0–2 years. Neuroimage, 2020. 218: p. 116946.

45. Dai, Y., et al., iBEAT: a toolbox for infant brain magnetic resonance image processing. Neuroinformatics, 2013. 11: p. 211–225.

46. Kim, H., et al., NEOCIVET: Towards accurate morphometry of neonatal gyrification and clinical applications in preterm newborns. Neuroimage, 2016. 138: p. 28–42.

47. Gruzieva, O., et al., Prenatal particulate air pollution and DNA methylation in newborns: an epigenome-wide meta-analysis. Environmental health perspectives, 2019. 127(5): p. 057012.

48. Neumann, A. and C. Cecil, Epigenetic timing effects on child developmental outcomes: A longitudinal meta-regression of findings from the Pregnancy and Childhood Epigenetics Consortium. In prep.

49. Walton, E., et al., Longitudinal epigenetic predictors of amygdala: hippocampus volume ratio. Journal of Child Psychology and Psychiatry, 2017. 58(12): p. 1341–1350.

50. Dunn, E.C., et al., Sensitive periods for the effect of childhood adversity on DNA methylation: results from a prospective, longitudinal study. Biological psychiatry, 2019. 85(10): p. 838–849.

51. Lussier, A.A., et al., Association between the timing of childhood adversity and epigenetic patterns across childhood and adolescence: findings from the Avon Longitudinal Study of Parents and Children (ALSPAC) prospective cohort. The Lancet Child & Adolescent Health, 2023.

52. Ray, P., S.S. Reddy, and T. Banerjee, Various dimension reduction techniques for high dimensional data analysis: a review. Artificial Intelligence Review, 2021. 54: p. 3473–3515.

53. Pearlson, G.D., J. Liu, and V.D. Calhoun, An introductory review of parallel independent component analysis (p-ICA) and a guide to applying p-ICA to genetic data and imaging phenotypes to identify disease-associated biological pathways and systems in common complex disorders. Frontiers in genetics, 2015. 6: p. 276.

54. Horvath, S. and K. Raj, DNA methylation-based biomarkers and the epigenetic clock theory of ageing. Nature reviews genetics, 2018. 19(6): p. 371–384.

55. Cole, J.H. and K. Franke, Predicting age using neuroimaging: innovative brain ageing biomarkers. Trends in neurosciences, 2017. 40(12): p. 681–690.

56. Nabais, M.F., et al., An overview of DNA methylation-derived trait score methods and applications. Genome Biology, 2023. 24(1): p. 28.

57. Wang, Y., et al., Polygenic prediction across populations is influenced by ancestry, genetic architecture, and methodology. Cell Genomics, 2023. 3(10).

58. Chen, J., et al., Pruning and thresholding approach for methylation risk scores in multi-ancestry populations. Epigenetics, 2023. 18(1): p. 2187172.

