## Supplementary for "Consortium Profile: The Methylation, Imaging and NeuroDevelopment (MIND) Consortium"

**Consortium Profile:**

**Words:** 4,968/5,000

**Tables and figures:** 2 tables, 3 figures

**Short title:** The MIND consortium

**Correspondence:**

Charlotte A.M. Cecil, Department of Child and Adolescent Psychiatry and Psychology, Erasmus MC University Medical Center Rotterdam, PO Box 2040, 3000 CA Rotterdam, the Netherlands. E-mail address:

**Supplementary materials**

**Study participants**

***Avon Longitudinal Study of Parents and Children***

The Avon Longitudinal Study of Parents and Children (ALSPAC) study is a prospective birth cohort, which invited pregnant women resident in Avon, UK with expected dates of delivery between 1st April 1991 and 31st December 1992 [1, 2]. The initial number of pregnancies enrolled was 14,541, of which 13,988 children who were alive at 1 year of age. When the oldest children were approximately age 7, efforts were made to bolster the sample size. The total sample size for analyses using any data collected after the age of seven is therefore 15,447 pregnancies, resulting in 15,658 foetuses. Of these 14,901 children were alive at 1 year of age. Participants with DNAm data were derived from the Accessible Resource for Integrated Epigenomic Studies (ARIES) [3], which initially included a subset of 1,018 mother-offspring pairs and was later extended.

Ethical approval for the study was obtained from the ALSPAC Ethics and Law Committee and the Local Research Ethics Committees Committees. Consent for biological samples has been collected in accordance with the Human Tissue Act (2004). Informed consent for the use of data collected via questionnaires and clinics was obtained from participants following the recommendations of the ALSPAC Ethics and Law Committee at the time. Please note that the study website contains details of all the data that is available through a fully searchable data dictionary and variable search tool: <http://www.bristol.ac.uk/alspac/researchers/our-data/>.

***Brazilian High Risk Cohort - Happy Mums***

The Brazilian High-Risk Cohort is a prospective high-risk cohort that commenced in 2010 and has been tracking children residing in Porto Alegre and São Paulo (Brazil) from mid-childhood onward. We used a two-stage design. We first assessed childhood symptoms and family history of psychiatric disorders in a screening interview, collecting information from 9,937 index children. In the second stage, a random subsample (intended to be representative of the community, n = 958) and a high-risk subsample (children at increased risk for mental disorders, based on family risk and childhood symptoms, n = 1554) were selected for further evaluation. From those, 750 children were invited to take part in a neuroimaging study and to provide blood samples for the assessment of peripheral blood biomarkers. These children, accompanied by their biological parents, participated in multiple research waves. The Scientific Committee and Research Ethical Commission of the Federal University of Rio Grande do Sul, and the Brazilian National Research Ethics Commission (CONEP), approved the protocol and informed consent form in accordance with the Brazilian and international regulatory framework under registration numbers IRB Nº: 20180018 and CAAE: 74563817.7.1001.5327.

***Drakenstein Child Health Study***

The DCHS is located in the periurban Drakenstein district, Western Cape, South Africa. Pregnant women were recruited while attending routine antenatal care at Mbekweni or TC Newman clinic between March 2012 and March 2015 [4-6]. Pregnant mothers were eligible for the study if they were 18 years or older, planned to attend antenatal care at one of the two clinics and intended to remain in the area for at least a year. Expecting mothers provided informed written consent at enrolment and were reconsented annually following childbirth. In total, 1137 mother–child dyads were enrolled in the study, of which four mothers had twins and one had triplets. Thus, 1143 children were enrolled in the study. The study was approved by the faculty of Health Sciences, Human Research Ethics Committee, University of Cape Town, Stellenbosch University and the Western Cape Provincial Health Research committee.

***Future of Families and Child Wellbeing Study***

The Future of Families and Child Wellbeing Study (FFCWS) is based on a stratified, multistage sample of 4,898 children born in large U.S. cities (population over 200,000) between 1998 and 2000, where births to unmarried mothers were oversampled by a ratio of 3 to 1. This sampling strategy resulted in the inclusion of a large number of Black, Hispanic, and low-income families. Mothers were interviewed shortly after birth and fathers were interviewed at the hospital or by phone. Follow-up interviews were conducted when children were approximately ages 1, 3, 5, 9, 15, and 22 (still being collected). When weighted, the data are representative of births in large US cities.

***Kids2Health – childhood***

The Kids2Health-Childhood study is a cohort of 535 children between 3-12 years of age in Berlin, Germany that was enriched for children with different types of chronic stress experiences (maltreatment, refugee stress, obesity as a metabolic stressor). The general design, research aims, and specific measurements of Kids2Health study have been approved by the Medical Ethical Committee of the Ludwig-Maximililans-Universität München. Written informed consent was obtained from the parents on behalf of the child.

***Kids2Health – infancy***

The Kids2Health-Infancy study is a cohort of 181 infants between 0-2 years of age in Berlin, Germany that was enriched for children whose mothers were exposed to childhood maltreatment. Pregnant mothers were recruited in early pregnancy and underwent a detailed clinical interview regarding childhood adverse experiences and were then followed up three times during pregnancy, at delivery and study visits with their offspring took place at 1, 6, 12 and 24 months age. The general design, research aims, and specific measurements of Kids2Health study have been approved by the Medical Ethical Committee of the Ludwig-Maximililans-Universität München. Written informed consent was obtained from the parents on behalf of the child.

***Michigan Twin Neurogenetic Study***

Participants in the Michigan Twins Neurogenetics Study (MTwiNS) were recruited from the Twin Study of Behavioral and Emotional Development in Children (TBED-C), part of the broader Michigan State University Twin Registry [7, 8]. The TBED-C identified twin families with children aged 6–10 years using birth records, focusing on families residing within 120 miles of East Lansing, Michigan. This region encompasses urban areas such as Detroit, Flint, and Lansing, along with suburban and rural locales. The overall sample comprised a population-based arm of 528 twin families and an "under-resourced" arm of 502 twin families from neighborhoods with above-average poverty levels, defined as over 10.5% of families living below the poverty line at the study's inception [9]. Detailed recruitment procedures are documented in [9].

Conducted in the USA, MTwiNS included typical neuroimaging exclusions, such as excluding individuals with metal in their bodies. The enrolment details for women and children are as previously mentioned. The study received ethics approval and informed consent from the Institutional Review Boards (IRB) at both Michigan State University and the University of Michigan.

***Oregon ADHD-1000***

The Oregon-ADHD-1000 is a community-recruited, longitudinal, case-control cohort of children (N=1483; age 7–11 years at baseline; 39% female) from northwest Oregon (USA) that is enriched for psychopathology. Details about study recruitment and data collection procedures have been published previously [10-12]. The local Institutional Review Board approved the studies. Parents provided written informed consent; children provided written informed assent. All families completed a multi-informant, multi-method screening process to establish eligibility and diagnostic group assignment for ADHD, non-ADHD, as well as comorbid disorders. The present study includes N=496 youth with good quality MRI and DNA methylation data.

***The CannTeen Study***

The CannTeen study is a purposively-recruited longitudinal cohort of adolescents (initial age 16-17 years) and adults (initial age 26-29 years) who either use cannabis regularly (at least weekly) or do not, based in London (UK). The initial sample comprised 274 participants, with 76 adolescents who use cannabis (50% girls), 63 adolescent controls (51% girls), 71 adults who use cannabis (47% women), and 64 adult controls (52% women). Exclusion criteria were: daily use of psychiatric medication, current treatment for a mental health disorder, personal history of a psychotic disorder, and regular use of any other illicit drug (apart from cannabis). Full cohort details can be found in Lawn, Mokrysz [13]. All participants gave a saliva sample for epigenetic analyses. 140 participants (n=35 in each group) completed MRI scans at baseline and one-year follow-up. The study received ethical approval from the University College London (UCL) ethics committee. As all participants were over 16 years of age, all participants gave their own informed consent to participate.

***The FinnBrain Birth Cohort Study***

FinnBrain Birth Cohort study is a prospective population-based cohort with recruitment around the city of Turku in Southwest Finland and the Åland island. Initial recruitment included 3808 mother-child dyads. Participants for the neonatal neuroimaging study were 97 healthy Finnish (Caucasian) neonates (57 males, 40 females) from 2 to 5 weeks of age, counted from the estimated due dat. All the infants were born full-term [between gestational weeks 37 and 42] and weighed more than 2500 g. The exclusion criteria for the infants were: occurrence of any perinatal complications with neurological consequences (e.g., hypoxia), scoring lower than 5 points in the 5 min Apgar, previously diagnosed CNS anomaly or an abnormal finding in a previous MRI scan. The families were contacted via telephone and eligibility for the study was assessed. After explaining the purpose and protocol of the study, a written informed consent was signed by the parents on behalf of each infant. The study was conducted according to the Declaration of Helsinki and was reviewed and approved by the Ethics Committee of the Hospital District of Southwest Finland (ETMK: 31/180/2011).

***The Generation R Study***

The Generation R Study is a prospective population-based cohort that follows children from fetal life onwards. Pregnant women living in the study area of Rotterdam, The Netherlands, with an expected delivery date between April 2002 and January 2006 were invited to participate [14]. A total of 9,778 mothers were enrolled. These mothers, their children and partners took part in several research waves. The general design, research aims, and specific measurements of The Generation R Study have been approved by the Medical Ethical Committee of Erasmus MC, in accordance with the Declaration of Helsinki of the World Medical Association. Written informed consent was obtained from the parents on behalf of the child. For data collected when the child was 12 years or older, the child also provided written informed consent.

***The Neuroimaging of the Children's Attention Project***

The Neuroimaging of the Children’s Attention Project (NICAP) is a longitudinal cohort examining brain and cognitive development in children with and without ADHD , with up to 3 repeated neuroimaging assessments between the ages 9-14. NICAP as an extension of the non-imaging longitudinal study the Children’s Attention Project recruited from the age 6-8years. Full cohort protocol details can be found in Silk, Genc [15] and Sciberras, Efron [16]. It is a community-based sample initially recruited from 43 socio-economically diverse primary schools across Melbourne, Australia. Exclusion criteria were: intellectual disability; previous known serious medical, neurological or genetic condition; moderate-severe sensory impairments; and insufficient English to participate. A total of 471 multimodal MRI scans were collected across three waves of data over five years. Being a community sample, females are well represented, representing roughly half the typical control sample and ~25% of the ADHD sample. This study was approved by The Royal Children’s Hospital (RCH) Human Research Ethics Committee (HREC #34071), and parents or guardians gave informed consent while participating children assent.

***Peri/postnatal Epigenetic Twins Study***

PETS is a prospective population-based cohort that follows twin children from mid-gestation onwards. A total of 25 women pregnant with twins were recruited half way through their second trimester (18-22 weeks) from three hospitals in Melbourne, Australia between 2007 and 2009 [17]. Data and multiple biosamples were collected from the mothers and their infants at four time-points during the pregnancy, at the time of the twins’ births, and again at 18 months, six years, and eleven years of age [18]. Buccal swabs were collected from children at each study wave. Ethical approval was granted by the Royal Children’s Hospital, Melbourne. Written informed consent was obtained from the parents and on behalf of children. Neurocognitive data, including cognitive assessment and brain imaging, were performed at 11 years (±3 months) [18]. Children with documented congenital brain malformations were exempt from MRI scans due to potential challenges in comparing these scans with those of unaffected children [18]. Additionally, participants were excluded from the MRI component if there were known contraindications, such as metal implants resulting from surgery. Individuals with severe neurosensory impairments were also excluded based on determinations by the Royal Children's Hospital Medical Imaging Team. All twins were invited to participate but MRI scans were only conducted on same-sex twins. Eighty-one twin pairs completed their cognitive assessment; 44 were monozygotic (MZ) and 37 dizygotic (DZ). Among the dizygotic twin pairs, 24 (65%) of them were of same sex. Fifty pairs completed their MRI scan, of which, 34 were MZ and 16 were DZ and of the same sex. Written informed consent was obtained from the parents on behalf of the child.

***UCIrvine Daily Experiences in Pregnancy Study***

The UCI cohort is a prospective, longitudinal study of ethnically and socio-demographically diverse pregnant women receiving prenatal care at the university and affiliated clinics, based in Irvine, Califiornia. All participants had singleton, intrauterine pregnancies, with no known cord, placental, or uterine anomalies, fetal congenital malformations, or presence of any conditions known to be associated with dysregulated neuroendocrine function or systemic corticosteroid medication use. The cohort comprised N = 253 mother-child dyads. All study procedures were approved by the university’s IRB, and all participants (pregnant women, and parents on behalf of their infants) provided written informed consent.

***Understanding Pregnancy Signals and Infant Development***

*Not available*

**DNA methylation**

***Avon Longitudinal Study of Parents and Children***

DNA methylation assays and data pre-processing was carried out as part of the Accessible Resources for Integrated Epigenomic Studies (ARIES, http://www.ariesepigenomics.org.uk/) project. ARIES is a sub-study of ALSPAC that initially included 1,018 mother-offspring pairs, selected based on availability of DNA samples. DNA was extracted from cord and peripheral blood samples (whole blood or buffy coat) using standard procedures and bisulfite-converted using the Zymo EZ DNA MethylationTM kit (Zymo, Irvine, CA). The methylation levels were assessed using the Illumina Infinium® HumanMethylation450K BeadChip assay (Illumina 450 K array) and Infinium MethylationEPIC BeadChip (Illumina, San Diego, CA, USA) across more than 485,000 and 850,000 CpG sites respectively, following the standard procedure. Illumina iScan was used to scan the arrays and initial quality evaluation was conducted using GenomeStudio (version 2011.1). Samples not meeting the QC criteria (with average probe *p*-value ≥0.01) were omitted from subsequent analysis and arranged for repeat assay. As an extra quality control step, genotype probes were cross-referenced with SNP-chip data of the corresponding individual to detect and eliminate any sample mismatches. For individuals lacking genome-wide SNP data, samples were marked if sex discrepancy was observed based on the methylation of the X-chromosome. Methylation level at each CpG site was denoted as a beta value (β), representing the ratio of the methylated probe intensity to the total intensity, ranging from 0 (no cytosine methylation) to 1 (complete cytosine methylation).

In total, there were 5,469 samples for five timepoints (in the children: birth=1,127; childhood=1,086; adolescence=1,073 (later extended to 2,857); in the mothers: pregnancy=1,100; middle aged mums=1,083) measured belonging to the 1,022 mother-child pairs. Sample quality control and functional normalization was completed using with *meffil* in R version 3.2.0. A total of 4,593 samples passed quality control (for detail see [19]). Samples were normalized using FN using *meffil*. ARIES was normalized using 10 control probe principal components derived from the technical probes informed by *meffil* scree plots. Slide and sample type effects in cord blood were not fully eliminated by normalization and further investigation showed that slide/plate effects were confounded by sample type in this dataset (e.g., because different sample types were not randomly distributed across slides and plates). Since then, samples in young adulthood (age 24) were added.

***Brazilian High Risk Cohort***

Each participant underwent the collection of four tubes of blood in a single session on the same day as the MRI scan. Two tubes were prepared with EDTA to facilitate DNA extraction, while the remaining two tubes were allocated for serum separation. This process yielded approximately 20 ml of venous blood per participant, with the tubes containing EDTA-preserved blood samples promptly stored at -20°C until DNA extraction procedures were conducted. DNA was isolated using a Gentra Puregene Kit (Qiagen) according to the manufacturer’s instructions. We identified 400 children with at least two neuroimaging data follow-up and DNA samples. We are planning to investigate DNAm in those 2 time points using the 953k Infinium MethylationEPIC v2.0 array (Illumina, San Diego, CA, USA). Subsequently, signal detection will be performed utilizing an iScan microarray scanner (Illumina), facilitating quantification and analysis of genome-wide DNA methylation levels and specificity. Furthermore, the necessary probes will be identified and selected for future exclusion from the analyses, considering factors such as quality, high background noise, cross-hybridization, or known artifacts, thereby ensuring the accuracy and reliability of the analysis.

***Drakenstein Child Health Study***

In the DCHS, umbilical cord blood was collected by trained staff after newborn delivery but before delivery of the placenta. Cord blood samples previously identified as being considerably contaminated with maternal blood (presumably resulting from mixing of blood during sample collection) Morin, Gatev [20], were excluded from the current analysis. As has been described fully previously [20]. Cord blood DNAm was measured using the Illumina Infinium HumanMethylation450K (Illumina, San Diego USA) as per manufacturer’s instructions. Raw data were imported into Illumina GenomeStudio Software for background subtraction and color correction and then exported for processing using the lumi package in R (version 3.2.3). Pre-processing and statistics were done using R 3.5.1. Raw iDat files were imported to RStudio where intensity values were converted into beta values. Background subtraction, color correction and normalization were performed using the preprocessFunnorm function. Batch effects were removed using ComBat from the R package sva. Cord blood cell type composition was predicted using the most recent cord blood reference data set and the IDOL algorithm and probe selection.

***Future of Families and Child Wellbeing Study***

At ages 9 and 15 salivary DNA was collected on 3,218 participants, and a random sample of those with both 9 and 15 samples were assay for DNA methylation. Approximately 500 ng of genomic DNA (quantified using the Quant-iT Picogreen ds DNA Assay Kit) was subjected to bisulfite conversion using the EZ-96 DNA Methylation Kit (Zymo Research) and analyzed with the Illumina Infinium Human Methylation450K (450K) or Illumina Infinium MethylationEPIC (EPIC) array according to the manufacturer’s protocol. Bisulfite conversion and array processing was performed by the Pennsylvania State College of Medicine Genome Sciences Core facility. Age 9 and 15 samples were run at the same time to minimize technical variation. Otherwise, samples were randomized. The red and green image pairs were read into R. QC of the methylation data was initially performed with EWAStools. Probes were removed if the detection value was greater than 0.01 or 0.05 for the 450K or EPIC arrays, respectively. Probes were also removed if the number of methylated or unmethylated bead count was fewer than four. Probes were also removed if they were identified by the ENmix function QCinfo using the default parameters. Samples were removed if they had outlier methylation or bisulfite conversion values, as identified by the ENmix QC function or if the sex predicted from the methylation data differed from the recorded sex. If the sequential samples from the same individual exhibited genetic discordance between visits the sample was flagged. The ENmix preprocessENmix and rcp functions were used to normalize dye bias, apply background correction and adjust for probe-type bias. Age of samples were confirmed using the Horvath Skin Blood epigenetic clock.

***Kids2Health – childhood***

Blood and saliva were sampled at timepoints T0 and T2 and DNA was extracted in a standardized and automated procedure with magnetic beads using a Chemagic 360 device (Revvity chemagen Technologie GmbH). DNA was bisulfite-converted using the EZ-96 DNA Methylation Kit (Zymo Research Corperation) and then run on the EPIC v2.0 Bead Chip (Illumina Inc., San Diego, USA). Methylation data were pre-processed in R according to a customized *minfi* [21] workflow (in-house pipeline: <https://github.molgen.mpg.de/mpip/EPIC_Preprocessing_Pipeline>). Data were normalized via stratified quantile normalization followed by beta-mixture quantile normalization (BMIQ [22]) and batch-corrected for plate, slide and array using *ComBat* via the *sva* R package [23]. We excluded probes on chromosomes X and Y, probes with mapping inaccuracies and flagged probes as reported by Illumina, and probes with a detection p-value > 0.01. For replicate probes, only the probe with the lowest detection p-value was retained. Samples were excluded in case of mean detection p-value > 0.05, mismatches of epigenetically predicted and phenotypic sex, or mismatches of genotypic and methylation data identified with MixupMapper [24]. Cell type proportions were estimated with the R package *epidish* [25] to use as covariates in statistical analyses.

***Kids2Health – infancy***

Cord blood and placenta were sampled at birth (T3). DNA was extracted from cord blood in a standardized and automated procedure with magnetic beads using a Chemagic 360 device (Revvity chemagen Technologie GmbH). Placenta DNA was extracted using an Illumina kit. DNA was bisulfite-converted using the EZ-96 DNA Methylation Kit (Zymo Research Corperation) and then run on the EPIC v2.0 Bead Chip (Illumina Inc., San Diego, USA). Methylation data were pre-processed in R according to a customized *minfi* [21] workflow (in-house pipeline: <https://github.molgen.mpg.de/mpip/EPIC_Preprocessing_Pipeline>). Data were normalized via stratified quantile normalization followed by beta-mixture quantile normalization (BMIQ [22]) and batch-corrected using *ComBat* via the *sva* R package [23]. We excluded probes on chromosomes X and Y, probes with mapping inaccuracies and flagged probes as reported by Illumina, and probes with a detection p-value > 0.01. For replicate probes, only the probe with the lowest detection p-value was retained. Samples were excluded in case of mean detection p-value > 0.05 and mismatches of epigenetically predicted and phenotypic sex. Additional cord blood samples were excluded in case of mismatches of genotypic and methylation data identified with MixupMapper [24] or a high likelihood of maternal blood contamination [20]. Cord blood cell type composition was estimated in *minfi* based on Gervin et al. [26]. Placenta cell types were estimated in a reference-based manner as described in Dieckmann et al. [27].

***Michigan Twin Neurogenetic Study***

A total of six samples were collected at four time points using two different tissues (dried blood spot (DBS) and saliva). Samples were collected at birth (DBS), Age 6-10 (Saliva), Early Adolescence (DBS and Saliva), and Mid Adolescence (about a18 months later; DBS and Saliva). Neonatal DBS were collected at birth via a heel prick and stored at the Michigan Neonatal Biobank until retrieval in 2023. Later DBS waves were collected via a Whatman 903 Protein saver card. Approximately 1/3 of a spot was used from the DBS samples. Saliva was collected using DNA Genotek Oragene 500 (age 6-10) and Oragene 600 (later waves). Approximately 300 ng of genomic DNA (quantified using the Quant-iT Picogreen) was subjected to bisulfite conversion using the EZ-96 DNA Methylation Kit (Zymo Research) and analyzed with the Illumina Infinium MethylationEPIC v2.0 (EPIC 2) array according to the manufacturer’s protocol. Bisulfite conversion and array processing was performed by University of Michigan Epigenetics Core. All 6 timepoints and each twin pair were run on the same plate to minimize technical variation. Otherwise, samples were randomized. The red and green image pairs were read into R. QC of the methylation data was initially performed with EWAStools. Probes were removed if the detection value was greater than 0.01. Probes were also removed if the number of methylated or unmethylated bead count was fewer than four. Probes were also removed if they were identified by the ENmix function QCinfo using the default parameters. Samples were removed if they had outlier methylation or bisulfite conversion values, as identified by the ENmix QC function or if the sex predicted from the methylation data differed from the recorded sex. If the sequential samples from the same individual (dizygotic twins) exhibited genetic discordance between visits the sample was flagged. The ENmix preprocessENmix and rcp functions were used to normalize dye bias, apply background correction and adjust for probe-type bias. Age of samples were confirmed using the Horvath Skin Blood epigenetic clock.

***Oregon ADHD-1000***

Genomic DNA was isolated from saliva, bisulfite converted, and assessed for DNA methylation on the MethylationEPIC BeadChip (Illumina, Inc.) using a standard protocol. Raw data were imported into Genome Studio v2011.1 (Illumina, Inc.) to investigate sample hybridization quality and to extract signal intensities for each probe. Data QC measures included: manual inspection of beta distributions, curation of control probes using the Illumina BeadArray Controls Reporter, manual inspection of total CpG intensity distributions, sex prediction, outlier sample detection, and comparison of SNP probes on the MethylationEPIC with genotypes, using the lumi and minfi R packages [21, 28]. Data was normalized using the dasen method in the wateRmelon R package [29].

Probes were removed for the following reasons: detection p-value < 0.01 in at least one sample, multi-mapping probes, probes which had an underlying SNP with a MAF of >= 0.01 in any examined subpopulation [30] or according to the Illumina manifest, and non-autosomal loci. In addition, outlier values were identified as those >4 times the interquartile range away from the sample mean for each probe, and were flagged as missing for subsequent analyses.

***The CannTeen Study***

At the baseline or second session (3 months) participants provided a single saliva sample (at least 4ml) in a GeneFiX saliva DNA collection tube. The samples were analysed by the King’s College London Genomics & Biomarker Core Facility. DNA methylation was measured using the Illumina Infinium MethylationEPIC (850K).

***The FinnBrain Birth Cohort Study***

Blood samples were acquired from infant’s umbilical cord blood at birth and DNA was extracted from both samples according to standard procedures at DNA unit of THL (Finnish Institute for Health and Welfare). Core blood DNAm was measured using the Infinium HumanMetylationEPIC BeadChip Infinium applying the manufacturer’s standard protocol at Erasmus MC, University Medical Center Rotterdam, Netherlands, corrected for background and followed by normalization with a subset quantile normalization approach (SWAN). The SWAN normalization process removed probes with detection p-value > 0.01 likewise probes with SNPs and cross-reactive probes and probes in X and Y chromosomes

***The Generation R Study***

DNA extracted at birth from cord blood, and at later ages using peripheral blood. DNA was extracted using the salting-out method and was then bisulfite-converted using the EZ-96 DNA Methylation Kit (Shallow) from Zymo Research Corporation. The DNA methylation analysis was conducted using two arrays: the Illumina Infinium HumanMethylation450 BeadChip (birth and later ages) and the MethylationEPIC v1.0 BeadChip (only at birth), both from Illumina Inc., San Diego, USA. The data underwent quality control procedures as prescribed by the CPACOR workflow, utilizing R software for data processing. All arrays were scrutinized for call rate, with those having a rate above 95% for the 450k array and 96% for the EPIC array being taken forward. Finally, we excluded samples with technical failures, such as failed bisulfite conversion, hybridization, or extension problems, as well as sex mismatch determined by the X and Y chromosome probe intensities.

***The Neuroimaging of the Children's Attention Project***

At each of the data collection wave, participants provided ~3mL saliva sample which were collected via passive drool into a 50 mL centrifuge tube and frozen at -30^o^C. Genomic DNA was extracted from saliva samples using a high salt extraction method. Briefly, 400 µL of saliva samples containing 20 µL of 20 mg/mL proteinase K (Promega, Madison, WI, USA) was added, and samples were vortexed to mix and centrifuged briefly, followed by incubation overnight at 55 °C. Following incubation, 400 µL of 5M NaCl (Sigma-Aldrich, NSW, Australia) was added to each tube. Tubes were vortexed and centrifuged at 9,000 g for 15 min at 4°C. Supernatants were then transferred to a fresh 1.5 mL microcentrifuge tube (Eppendorf South Pacific) and the pellet was discarded. To each tube, 1 mL of 100 % ethanol was added, and the tube was mixed by inversion six times and centrifuged at 18,000 g for 5 min at room temperature. The supernatant was discarded, wash step was repeated, and the pellet was left to air dry at room temperature. Pellets were resuspended 40 µL 1 x 10mM Tris-HCl containing 1mM EDTA•Na_2_ (Astral Scientific, NSW, Australia). DNA concentration and quality was measured by fluorometry (Qubit, Thermo Fisher Scientific, VIC, Australia) and spectrophotometry (NanoDrop, Wilmington, DE, USA). DNA samples were bisulphite treated and hybridised to Infinium Human Methylation 850 BeadChip arrays (Illumina, San Diego, CA, USA)

***Peri/postnatal Epigenetic Twins Study***

DNA was extracted using the salting-out method Ollikainen, Smith [31] from 1288 buccal swabs across the four study waves. DNA methylation analysis was conducted following bisulphite conversion using the MethylationEPIC v1.0 BeadChip from Illumina Inc., San Diego, USA. Samples from the same twin pair were located on the same array where possible and pairs were randomised across arrays. Analysis of DNA methylation data was performed using the R programming language. Initially, sample quality control measures were implemented by eliminating samples that do not meet the signal to noise ratio (detection p-value > 0.01). Subsequently, poor-performing probes failing to meet specified quality parameters, with a detection p-value exceeding 0.01, were systematically excluded. Further refinement included the removal of control probes, those targeting CpGs with known SNPs, and probes situated on sex chromosomes to minimize sex-dependent variability [32].

***UCIrvine Daily Experiences in Pregnancy Study***

Genomic DNA was isolated from neonatal heel stick blood samples collected shortly after birth (N=129). Genomic DNA (500ng) was bisulfite converted using the EZ DNA methylation kit (Zymo Research, Irvine, CA, USA) and genome-wide DNA methylation described using the Infinium human MethylationEPIC Beadchip (Illumina). Signal extraction from raw data files (.idat), quality control, and normalization were performed using Meffil in R. We used functional normalization to address technical variation. Four samples were removed due to missing information on biological sex (n=3) or low signal intensity (n=1). DNA methylation data were processed in R using the Minfi package (Aryee et al., 2014). All samples passed standard Minfi quality control (QC) (QC threshold=10.5) (Aryee et al., 2014), and had a high call rate (>95%). Probes with a low call rate (<75%), a high detection p-value (p>0.0001) (Lehne et al., 2015) and a low number of beads (less than 3 in >5% of the cohort) were removed. Predicted sex (from DNA methylation of the sex chromosomes) (Aryee et al., 2014) and reported sex for all participants was consistent for all samples. Non-specific probes or probes with SNP-disrupting polymorphisms were removed (Chen et al., 2013; Price et al., 2013; McCartney et al. 2016), along with all the probes on the sex-chromosomes. Unwanted technical variation was accounted for, in part, using functional normalization based on principal component analysis of control probes on the 850K (Fortin et al., 2014).

Proportions of blood cell-types were estimated through a deconvolution approach validated for use with neonatal blood samples [33]. DNA methylation data and paired neuroimaging phenotypes were available on 82 participants.

***Understanding Pregnancy Signals and Infant Development***

*Not available*

**Image acquisition and pre-processing**

***Avon Longitudinal Study of Parents and Children***

As described in Sharp et al., [34], subset of ALSPAC offspring took part in three different neuroimaging studies: the ALSPAC Testosterone study (n=513), the ALSPAC Psychotic Experiences (PE) study (n=252), and the ALSPAC Schizophrenia Recall-by-Genotype (SCZ-RbG) study (n=196), with some participant overlap (unique participant n before QC = 889). All data were obtained at Cardiff University Brain Research Imaging Centre on General Electric 3T HDx scanner with an 8-channel head coil. When possible, scanning protocols were aligned across sub-studies.

**ALSPAC-Testosterone Study*.*** Structural coronal T1 images were acquired with the following imaging parameters: 3D fast spoiled gradient echo (FSPGR) with 168–182 oblique-axial AC-PC slices, 1 mm isotropic resolution; flip angle of 20°; repetition time (TR) of 7.9 ms; echo time (TE) of 3.0 ms; inverse time (TI) of 450 ms; 1mm × 1mm x 1mm voxel size; slice thickness 1 mm; field of view (FOV) 256 × 192 mm matrix. Each T 1-weighted scan took about 7.15 minutes. Diffusion Tensor Imaging (DTI) data were acquired with a dual spin-echo, single shot echo-planar imaging sequence. This included 30 gradient orientations and 3 non-diffusion weighted images (b = 0 s/mm 2), which were obtained using the following parameters: resolution of 2.4 × 2.4 × 2.4 mm; FOV of 230 × 230 mm; acquisition matrix of 96 × 96; slice thickness of 2.4mm; number of slices = 60 (oblique-axial AC-PC); TR/TE of cardiac gated/87ms (effective); b = 1200 s/mm 2, T_1_ = 0; flip angle of 90°; number of excitations (NEX) = 1; parallel imaging acceleration factor (ASSET) = 2;30. The acquisition duration ranged from 15 to 20 minutes. For task-based fMRI (dynamic faces paradigm) the acquisition parameters were as follows: GE-EPI (AC-PC); resolution of 3.4 × 3.4 mm; FOV of 220 × 220 mm; matrix of 64 × 64; slice thickness of 2.4 mm; number of slices of 45; gap of 1 mm; TR/TE of 3000/35; TI of 0; flip angle of 90; NEX of 1, acquisition time being 6.42. The order in which the slices were acquired alternated in an ascending pattern. Additional modalities included Multi-component driven equilibrium single-pulse observation of T1 and T2 (mcDESPOT) and Magnetization transfer MRI (see Sharp, McBride [34]).

**ALSPAC Psychotic Experiences (ALSPAC-PE).** T1-weighted structural images with a 1mm isotropic resolution were acquired using a FSPGR sequence (TR = 7.8 ms, TE = 3.0 ms, TI = 450 ms, flip angle = 20°, acquisition matrix = 256 × 192, zero-padded matrix = 256 × 256). T2-weighted whole brain scans were acquired using a coronal TSE sequence with the following parameters: TR=10000 ms for 3T, TR=9000 ms for 1.5T; TE=14 ms for 3T, TE=64 ms for 1.5T; FA=149 for 3T, FA=180 for 1.5T; Bandwidth=193 for 3T, Bandwidth=149 for 1.5T; voxel size same as for T 1 scans; NEX=1 for the 3T, NEX=2 for the 1.5T. Acquisition of each volume took approximately 14.5 minutes. Diffusion MRI comprising a cardiac-gated diffusion-weighted spin-echo echo-planar imaging sequence was used to obtain high angular resolution diffusion weighted images (HARDI). A total of 60 gradient orientations and 6 unweighted (b = 0 s/mm 2) images were acquired with the following parameters: TR = cardiac-gated, TE = 87 ms, acquisition matrix = 96 × 96, zero-padded matrix = 128 × 128), FoV = 230 × 230 mm. Following zero-padding, the reconstructed image resolution for the HARDI scans was 1.8 × 1.8 × 2.4 mm. The acquisition parameters for task-based fMRI (N-Back working memory paradigm) were as follows: T2*-weighted gradient-echo echo-planar images along the axial plane parallel to the AC–PC line (TR = 2000 ms, TE = 30ms, flip angle = 75°, FOV = 240 × 240mm, resolution = 3.75 × 3.75 × 3.5 mm). mcDESPOT was also acquired (see Sharp, McBride [34]).

**ALSPAC-Schizophrenia Recall-by-Genotype*.*** High-resolution 3-dimensional T 1-weighted images were acquired using a 3D FSPGR with contiguous sagittal slices of 1 mm thickness (TR=7.9 s, TE = 3.0 ms, TI = 450 ms, flip angle = 20°, FOV = 256 × 256 × 176 mm to yield 1 mm 3 isotropic voxel resolution images) were collected for each participant. T1-weighted structural scans were acquired using an oblique axial, 3D FSPGR with the following parameters: TR = 7.9 ms, TE = 3.0 ms, inversion time = 450 ms, flip angle = 20°, 1 mm isotropic resolution, with a total acquisition time of approximately 7 minutes. HARDI data were acquired using a cardiac-gated, peripherally gated twice-refocused spin-echo EPI sequence. A total of 60 gradient orientations and three non-diffusion weighted (b = 0 s/mm 2) images were acquired with effective TR/TE of 15R-R intervals/87ms, FoV = 230 × 230 mm, acquisition matrix = 96 × 96, zero-padded matrix = 128 × 128. Following zero-padding, the reconstructed image resolution for the HARDI scans was 1.8 × 1.8 × 2.4 mm. Sets of 60 contiguous 2.4-mm thick axial slices were obtained, with diffusion-sensitizing gradients applied along 30 isotropically distributed gradient directions (b = 1,200 s/mm 2). For task-based (N-Back working memory task, reversal learning) fMRI, gradient echoplanar imaging data were acquired for each participant using the following parameters: 35 slices, slice thickness = 3 mm/1 mm gap, acquisition matrix = 64 × 64; FOV = 220 mm, TR = 2000 ms, TE = 35 ms, flip angle = 90°, ASSET factor; 2. All functional images were first motion scrubbed, where TRs with a frame wise displacement of greater than 0.9 were removed. A total of 354 volumes (12 minutes) for the reversal learning and 265 volumes (9 minutes) for the N-Back study were acquired. Resting-state and task-based (N-Back, visual stimulation, mismatch negativity) Magnetoencephalography (MEG) data were also acquired (see Sharp, McBride [34]).

Please see Table 2 in Sharp, McBride [34] for the number of participants available across ALSPAC sub-studies, modalities, and tasks.

**Extraction of image-derived phenotypes from structural MRIs.** T1-weighted images were processed using the automated FreeSurfer brain imaging software package (Version 6.0.0) via the ‘recon-all’ command including the -qcache flag. Processing includes an automated pipeline of removal of non-brain tissue, voxel intensity correction for B 1 field inhomogeneities, segmentation of voxels into white matter, grey matter or cerebral spinal fluid, and generation of surface-based models of white and grey matter. Reconstructed images were subjected to QC steps following the ENIGMA consortium structural image processing protocol.

***Brazilian High Risk Cohort***

Resting-state functional neuroimaging data were acquired using two 1.5 T MRI GE scanners (Signa HDX in São Paulo and Porto Alegre, Brazil) with identical acquisition parameters. Each participant underwent the acquisition of one hundred and eighty whole-brain EPI volumes (TR = 2000 ms, TE = 30 ms, slice thickness = 4 mm, gap = 0.5 mm, flip angle = 80°, matrix size = 80 × 80, reconstruction matrix = 128 × 128, voxel size = 1.875 × 1.875 mm, NEX = 1, slices = 26, total acquisition time of 6 min). Participants were instructed to keep their eyes open and fixate gaze at a painted target. T1-weighted scans (3D FSPGR sequence) were obtained with up to 160 axial slices (TR = 10.91 ms, TE = in phase 4.2 ms, thickness = 1.2 mm, flip angle = 15°, matrix size = 256 × 192, FOV = 24.0 × 18.0 cm, NEX = 1). Diffusion Weighted Imaging was acquired using the same parameters as the previous scans, with a sequence duration of approximately 6 minutes and 20 seconds, including 16 directions (including b0), TR = 11,600 ms, TE = 90 ms, matrix size = 256 × 256, slice thickness = 3 mm, angle = 90, and b-value = 800 s/mm2.

The images were processed using scripts based on routines from AFNI, FSL, and Freesurfer. Data of each participant underwent preprocessing steps including discarding the first four volumes, head motion correction, skull-stripping and despiking, linear detrending, spatial smoothing (Gaussian kernel, FWHM = 6 mm), and grand-mean scaling. Fractional amplitude of low-frequency fluctuations (fALFF) was calculated for each intracranial voxel in the frequency range from 0.01 to 0.1 Hz. The fALFF maps were normalised to z-scores considering the whole brain and then spatially normalised to standard space using a non-linear transform, the individual T1 image, and the Montreal Neurological Institute (MNI152) template.

***Drakenstein Child Health Study***

Sagittal T2-weighted MR images were acquired on a 3T Siemens Magnetom Allegra scanner (Erlangen, Germany) with scan parameters: TR = 3500ms; TE = 354 ms; FOV = 160 x 160 mm; 128 slices; voxel size = 1.3 x 1.3 x 1.0 mm. The sequence took 5 min. 41 s. to acquire. Images were brain-extracted using FSL v5.0 and pre-processed further using Statistical Parametric Mapping software (SPM8) run in Matlab R2017B. Images were registered and normalised to the University of North Carolina (UNC) neonate T2 template [35] and segmented into grey matter, white matter and cerebrospinal fluid according to the UNC neonate probabilistic maps. Segmentation accuracy was confirmed through visual inspection prior to analysis. Left and right thalamus, caudate, putamen, pallidum, hippocampus and amygdala volumes were extracted as regions of interest defined in the automated anatomical labelling atlas (AAL; [36]), adapted for neonates [35].

***Future of Families and Child Wellbeing Study***

The neuroimaging subsample of the FFCWS study is called the Study of Adolescent to Adult Neurodevelopment or SAND. A cohort of 237 families from midwestern sites (*N* = 237; mean age 15.87 years; 52% females, 76% Black). Magnetic Resonance Imaging (MRI) scans were acquired using 3 T GE Discovery MR750 scanner with 8-channel head coil at the University of Michigan Functional MRI laboratory. Head movement was limited through the use of head paddings and detailed instructions provided to participants. T1-weighted gradient echo images were first captured (TR = 12 ms, TE = 5 ms, TI = 500 ms, flip angle = 15°, field of view = 26 cm, slice thickness=1.44 mm, 256 × 192 matrix, 110 slices). [Diffusion MRI](https://www.sciencedirect.com/topics/neuroscience/diffusion-mri) (dMRI) data were then acquired using spin-echo EPI diffusion sequence using repetition time of 7250 ms, minimum echo time, 128 × 128 acquisition matrix, FOV = 22 cm, 3 mm no-gap thick slices with 40 slices acquired using alternating-increasing order, b-value of 1000 s/mm2, 64 non-linear directions. dMRI images were first inspected visually for quality, and slices with an average intensity < 4 standard deviations or more were marked as outliers and replaced with predicted models. Participants were excluded if more than 5% of slides were replaced and images for 10 participants who had most replaced slices were further visually inspected.

***Kids2Health – childhood***

Magnetic resonance imaging (MRI) of the brain was acquired on a 3T Siemens Fit, Tim+Dot (Siemens Healthineers, Erlangen, Germany) using a 64 channel head coil. The MRI protocol comprised T1 weighted (T1w), T2 weighted (T2w), Resting State as well as task based functional imaging and diffusion tensor imaging (DTI). Three-dimensional T1w MPRAGE and T2w SPACE images were acquired for all participants using the following scan parameters: T1w) TE = 2.22 ms, TR = 2400 ms, flip angle = 8°, bandwidth = 220 Hz/Px, FOV = 256 mm, resolution 0.8 mm isotrope, TA = 6:38 min. T2w) TE = 563 ms, TR = 3200 ms, bandwidth = 744 Hz/Px, FOV = 256 mm, resolution 0.8 mm isotrope, TA = 5:57 min. Volumetric segmentations as well as cortical parcellations and transformation to cifti-space were carried out using the anatomical part of the ABCD-HCP-pipeline [37](https://github.com/DCAN-Labs/abcd-hcp-pipeline, OHSU, Portland Oregon & MIDB, Minneapolis Minnesota). In brief, this pipeline normalizes the brain data entailing skull stripping, denoting and bias field correction using T1w and T2w data. Within the pipeline Freesurfer [38] and ANTs [39] are used to reconstruct the cortical surfaces (pial and white matter surface) from the normalized data as well as the anatomical segmentation and the subsequent transformation to a standard surface template representing children.

The DTI data were acquired with an axial spin echo, by using an echo-planar imaging sequence with six b = 0 volumes and 74 diffusion volumes in each encoding direction, AP and PA (TR = 3222 ms, TE = 89.2 ms, FOV = 210 mm × 210 mm, acquisition matrix = 140 × 140, slice thickness = 1.5 mm, voxel size = 1.5 mm × 1.5 mm x 1.5 mm, number of slices = 92, asset acceleration factor = 4, b = 1500 s/mm2 and b = 3000 s/mm2). DTI images were pre-processed through the DMRIprep pipeline [40]. This pipeline compromises a slice wise checking as well as an interlace checking, susceptibility correction and Eddy current correction using FSL [41]. Brain gets skull stripped and a full brain tractography is performed for QC purposes. Subsequently, the tensor model gets estimated.

For resting state as well as task based fMRI, echo planar imaging was used (TR = 800 ms, TE = 37 ms, flip angle = 52°, matrix = 104 × 104, FOV = 208 mm × 208 mm, slice thickness = 2 mm). Resting state was acquired in two runs, each 5:46 minutes leading to 840 volumes in total. Data were pre-processed using the functional part of the ABCD-HCP-pipeline. This part comprises a distortion correction using reverse phase encoding spin echo field maps with the same scanning parameters as stated above. All data has been normalized and corrected for head movements as well as registered to a standard children template by projecting the normalized functional data onto the template surface. Additionally, a time domain filter was used to filter respiratory artifacts from the bold data.

***Kids2Health – infancy***

Brain MRI data from the infant cohort was acquired on the very same Siemens 3T scanner with a 64 channel head coil, where the neck elements were switched on additionally to adjust for the position of the infant inside the coil. The MRI protocol contained T1w, T2w, resting state functional imaging as well as DTI. T1w and T2w sequences were scanned in the exact same way as described above to not introduce any changes from infancy to toddlerhood. Volumetric segmentations were carried out using the BIBSnet pipeline [42]
(<https://bibsnet.readthedocs.io/en/latest/>), where in its core a deep learning nnU-Net assigns the tissue labels according to the Desikan atlas. Atlas refinement as well as registering to a surface-template-space is achieved by the nibabies [43] and xcp-d [44] pipelines.

DTI data was acquired and processed the exact same way as described for the childhood cohort.

Resting State functional imaging was acquired as described for the childhood cohort. Data preprocessing was carried out using nibabies and xcp-d to normalize and denoise the images as well as correct for distortion using everse phase encoding spin echo field maps. The data has been corrected for head movement and filtered from respiratory artifacts and finally mapped onto a infant surface template.

***Michigan Twin Neurogenetic Study***

MRI scans were acquired using a General Electric Discovery MR750 3T scanner, encompassing task-based fMRI (emotional faces, simple reward processing, Go/No-Go), resting state fMRI, diffusion MRI (dMRI), and structural MRI (sMRI). To enhance MRI data acquisition and align our protocol with the Adolescent Brain Cognitive Development Study (Casey et al., 2018), we revised our acquisition protocol after scanning the first 140 families (280 twins). For these initial families, each participant underwent one run of 298 volumes using BOLD functional images acquired with an 8-channel head coil and a reverse spiral sequence (TR/TE = 2000/30 milliseconds, flip angle = 90°, FOV = 22 cm), covering 43 interleaved oblique slices of 3-mm thickness. High-resolution T1-weighted SPGR images (156, 1 mm-thick slices) were aligned with the AC-PC plane and used for normalizing functional images. For the subsequent 214 families (428 twins), each participant had one run of 730 volumes, with BOLD functional images acquired via a 32-channel head coil and a gradient-echo sequence with multiband acquisition (TR/TE = 800/30 milliseconds, flip angle = 52°, FOV = 21.6 cm), covering 742 interleaved axial slices of 2.4-mm thickness. High-resolution T1-weighted SPGR images (208, 1 mm-thick slices) were aligned with the AC-PC plane and used for normalizing the functional images. For both acquisition sequences, BOLD functional images encompassed the entire cerebrum and most of the cerebellum to maximize coverage of limbic structures. To take advantage of improvements in MRI data acquisition and harmonize our protocol with the Adolescent Brain Cognitive Development Study (Casey et al., 2018), we altered our acquisition protocol after the first 140 families (280 twins). For the first 140 families, high-resolution T1-weighted SPGR images were acquired via an 8-channel head coil (156, 1 mm-thick slices, TI = 500, flip angle = 15°, FOV = 25.6 mm²). For the subsequent 214 families (428 twins), high-resolution T1-weighted SPGR images were acquired via a 32-channel head coil (208, 1 mm-thick slices, TI = 1060, flip angle = 8°, FOV = 25.6 mm²). We purposefully kept the slices the same thickness to allow for better harmonization of the two acquisition sequences. Functional MRI data for both acquisition sequences were preprocessed and analyzed using Statistical Parametric Mapping version 12 (SPM12; Wellcome Trust Centre, London, United Kingdom), with postprocessing control for artifacts using the Artifact Detection Tools (ART) software package (http://www.nitrc.org/projects/artifact_detect/). Participants with low amygdala coverage (<90% signal coverage), low task performance (<70% accuracy), and >5% motion outliers identified using ART were excluded from analyses. T1-weighted scans were processed using the freely available and extensively validated FreeSurfer software (version 6.0.0, http://surfer.nmr.mgh.harvard.edu/). The FreeSurfer automated stream consists of several processing steps including skull stripping, Talairach transformation, subcortical structure labeling, surface extraction, spherical registration, and cortical parcellation. The data for each participant were resampled to an average participant and surface smoothing was performed using the qcache command and a 10-mm full-width half-maximum Gaussian kernel prior to statistical analysis. All FreeSurfer output underwent quality checking according to the protocol established by the ENIGMA workgroup (see https://enigma.ini.usc.edu/), which included visual inspection and outlier detection.

***Oregon ADHD-1000***

Structural and functional data were acquired using a 3T Siemens Tim TRIO equipped with a 12-channel head coil. 580 subjects completed T1-weighted structural imaging (TR = 2300ms, TE = 3.58ms, FOV = 256 x 256, orientation = sagittal) and passed quality control. Voxel size was either 1mm x 1 mm with 1.1mm slice thickness (N = 460) or 1mm isotropic (N = 120).

Processing of structural images was carried out using FreeSurfer (v. 6.0.0, http://surfer.nmr.mgh.harvard.edu/) cross-sectional procedures and included: skull stripping, intensity normalization, gray-white matter segmentation, surface reconstruction, and volumetric segmentation and labeling of cortical and sub-cortical brain structures. Visual quality control was carried out via methods similar to those described for the Adolescent Brain Cognitive Development (ABCD) Study [45]. Briefly, the accuracy of the cortical surface reconstruction was reviewed for each subject, and subjects were excluded if there was severe motion, intensity inhomogeneity, white matter underestimation, pial overestimation, or magnetic susceptibility artifact present.

Resting state data was acquired in three 5-minute blocks using blood oxygen level-dependent (BOLD) contrast (TR = 2500ms, TE = 30ms, flip angle = 90 degrees, FOV = 240mm, in-plane resolution = 3.8mm x 3.8mm with a slice thickness of 3.8 mm), and full details can be found in Nigg, Karalunas [12].

Diffusion‐weighted imaging scans were collected using a whole‐brain, high‐angular resolution, echo‐planer imaging sequence (repetition time = 9100 ms, echo time = 88 ms, field of view = 256 mm^2^, slices = 72, slice thickness = 2 mm) with gradient‐encoding pulses in 30 directions (b‐value = 1000 s/mm^2^) and six images collected with a b‐value of 0 s/mm^2^. Participants received either two (scan time = 11:24) or three (scan time = 16:52) DWI runs. Data were processed via the FMRIB Software Library Diffusion Toolbox (v 5.0.11). Details of the quality control procedures are described in Jones, Nagel [46].

***The CannTeen Study***

We acquired data on a 3-T Siemens Verio MRI scanner. Scanning parameters for each of the scans can be found in the table below. Structural images were acquired using an MPRAGE sequence. Functional images were acquired using a ‘multiband’ or Simultaneous Multi-Slice (SMS) Echo-Planar Imaging (EPI) sequence, based on the multiband EPI WIP v012b provided by the University of Minnesota (Xu et al., 2013). fMRI data have been pre-processed and analysed using FSL [47].

**Table 1.** CannTeen MRI scanning parameters

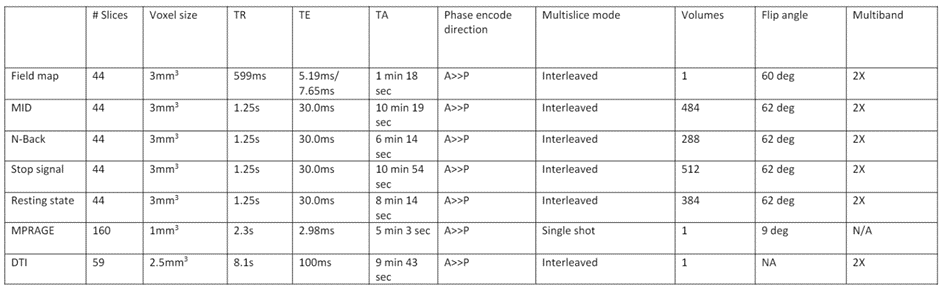

***The FinnBrain Birth Cohort Study***

Participants were imaged with a Siemens Magnetom Verio 3T scanner (Siemens Medical Solutions, Erlangen, Germany). The imaging protocol was 40-60 min long and included an axial PD-T2-TSE (Dual-Echo Turbo Spin Echo) sequence, a sagittal 3D-T1 (T1-weighted MPRAGE) sequence, and diffusion tensor imaging (DTI) sequence. The slice thickness of the PD-T2 sequence was only 1 mm, to achieve an isotropic voxel size of 1.0 mm3. TR (time repetition) time of 12,070 ms and effective TE (time echo) times of 13 ms and 102 ms were used to produce both PD-weighted and T2-weighted images from the same acquisition. The total number of slices was 128. Sequence parameters were optimized so that “whisper” gradient mode could be used in PD-T2 TSE and 3D-T1 sequences to reduce acoustic noise during the scan. All brain images were assessed for incidental findings by a pediatric neuroradiologist.

The volumes of the left and right amygdalae and hippocampi were assessed for each subject via label-fusion-based methods. These methods depend on achieving good registrations between the subjects and the template. This is increasingly difficult to achieve the further the template is from the subjects in terms of similarity. Thus, we constructed a template based on the subjects in this study. We then manually labeled the subcortical structures on this template, and constructed a library of warped versions of the labeled template such that the library represented the morphological variation in the sample. We then labeled the individual brains via label-fusion-based methods, and calculated the volume of each structure. The details of this approach are described in our prior article [48].

***The Generation R Study***

Brain magnetic resonance imaging was acquired on a 3-T GE MR750W scanner (General Electric, Milwaukee, WI), with an eight-channel head coil. As part of a larger scanning protocol, the Generation R Study acquired T1, DTI and resting state fMRI scans (White et al., 2018). T1-weighted scans were acquired using an inversion recovery fast spoiled gradient recalled echo sequence (TR = 8.77 ms, TE = 3.4ms, TI = 600ms, flip angle = 10°, acquisition matrix = 220 x 220, FOV = 220 mm x 220 mm, slice thickness = 1 mm, number of slices = 230, voxel size = 1mm x 1mm x 1mm, ARC Acceleration = 2). Automatic volumetric segmentations of the T_1_-weighted images were carried out with the FreeSurfer analysis suite, version 6.0 (Fischl et al., 2004). The DTI data were acquired with an axial spin echo, by using an echo-planar imaging sequence with three b = 0 volumes and 35 diffusion volumes (TR = 12,500 ms, TE = 72.8 ms, FOV = 240 mm × 240 mm, acquisition matrix = 120 × 120, slice thickness = 2 mm, voxel size = 2 mm × 2 mm x 2 mm, number of slices = 65, asset acceleration factor = 2, b = 900 s/mm^2^). DTI images were pre-processed through the FMRIB Software Library (FSL) (v 6.0.2).  For resting state fMRI, echo planar imaging was used (TR = 2,000 ms, TE = 30 ms, flip angle = 85°, matrix = 64 × 64, FOV = 230 mm × 230 mm, slice thickness = 4 mm). Data were pre-processed using the Functional MRI of the Brain (FMRIB) Software Library. A full description of the image processing pipeline and quality assurance can be found elsewhere [49].

***The Neuroimaging of the Children's Attention Project***

Multimodal neuroimaging data was collected from a 3-Tesla Siemens MRI scanner at a single-site. Prior to scanning all children participated in a 30-minute mock MRI training session. A multimodal imaging protocol included T1 weighted MEMPRAGE, T2 weighted, multishell Diffusion Weighted Imaging (b=2800, 2000, and 1000) and Resting State Functional MRI. A full description of the imaging protocol can be found in Silk, Genc [15].

***Peri/postnatal Epigenetic Twins Study***

A paediatric neuroimaging protocol (45mins) was created for this study, optimised for the collection of estimates of brain structure and morphology, white matter connectivity and spontaneous intrinsic neural activity (BOLD) [18]. Brain magnetic resonance imaging (MRI) was acquired on a dedicated 3-Tesla MAGNETOM Prisma system (Siemens Medical Systems, Erlangen, Germany) equipped with a 32-channel phased-array head coil. T1-weighted imaging data were acquired using an inversion recovery fast spoiled gradient recalled echo sequence (TR = 2550 ms, TE = 3.94 ms, TI = 1550 ms, flip angle = 8°, acquisition matrix = 640 x 640, FoV = 256 mm x 256 mm, slice thickness = 1 mm, number of slices = 208, voxel size = 0.4 mm x 0.4 mm x 0.9 mm, echo spacing = 9.5 mm).
Diffusion weighted data was acquired in a multi-shell protocol (b = 3000, 2000, 1000 s/mm^2^ + interleaved b = 0 s/mm2) in an anterior-posterior phase encoding direction, (TR = 3,500 ms, TE = 74.4 ms, FoV = 220 mm × 220 mm, acquisition matrix = 110 × 110, slice thickness = 2 mm, voxel size = 2 mm × 2 mm x 2 mm, number of slices = 90, echo spacing = 0.53 mm). In addition two additional reverse phase encoding scans (b= 0 volumes) were along the posterior-anterior direction to correct for susceptibility-induced geometric distortion. Resting state fMRI data was acquired using multiband echo planar imaging (5mins46sec duration, MB=3, TR = 800 ms, TE = 37 ms, flip angle = 52°, FoV = 208 mm × 208 mm, voxel size = 2 mm × 2 mm x 2 mm, matrix = 104 × 104, slice thickness = 4 mm, number of slices = 72, echo spacing = 0.58 mm).

***UCIrvine Daily Experiences in Pregnancy Study***

Magnetic resonance imaging (MRI) was performed in unsedated newborns during natural sleep using a Siemens 3T scanner (TIM Trio, Siemens Medical System Inc., Germany). T1-weighted images were obtained using a three-dimensional (3D) magnetization prepared rapid gradient echo (MP-RAGE) sequence (TR 2400 ms; TE 3.16 ms; TI 1200 ms; Flip Angle 8°; 6:18 minutes) and T2-weighted images were obtained with a turbo spin echo sequence (TR 3200 ms; TE1 13 ms; TE2 135 ms; Flip Angle 180°; 4:18 minutes). The spatial resolution was a 1 × 1 × 1 mm voxel for T1-weighted images and 1 × 1 × 1mm voxel with 0.5 mm interslice gap for T2-weighted images.

In N = 114 neonates an attempt was made to acquire an MRI scan; the complete MRI sequence was obtained in N = 94 neonates. Scan acquisition was not attempted in N = 17 infants because they did not fall asleep during the MRI study visit. Image quality control (QC) feedback was provided using a four point scale (1–4) developed for infant scanning, and based on a widely used visual QC protocol [50]. Criteria for exclusion was a QC score of 4 (N = 6 excluded), representing subjects with artifact contamination to a degree that renders image processing unreliable. A further N = 2 had to be excluded due to significant abnormalities as reviewed by a clinical neuroradiologist.

Tissue segmentation was performed using a multi-atlas based iterative expectation maximization segmentation algorithm as previously described [51, 52]. Brain tissue was classified as gray matter, white matter, and cerebrospinal fluid.

***Understanding Pregnancy Signals and Infant Development***

*Not available*

**References**

1.	Boyd, A., et al., *Cohort profile: the ‘children of the 90s’—the index offspring of the Avon Longitudinal Study of Parents and Children.* International journal of epidemiology, 2013. **42**(1): p. 111-127.

2.	Fraser, A., et al., *Cohort profile: the Avon Longitudinal Study of Parents and Children: ALSPAC mothers cohort.* International journal of epidemiology, 2013. **42**(1): p. 97-110.

3.	Relton, C.L., et al., *Data resource profile: accessible resource for integrated epigenomic studies (ARIES).* International journal of epidemiology, 2015. **44**(4): p. 1181-1190.

4.	Donald, K.A., et al., *Drakenstein Child Health Study (DCHS): investigating determinants of early child development and cognition.* BMJ paediatrics open, 2018. **2**(1).

5.	Stein, D.J., et al., *Investigating the psychosocial determinants of child health in Africa: The Drakenstein Child Health Study.* Journal of neuroscience methods, 2015. **252**: p. 27-35.

6.	Zar, H.J., et al., *Investigating the early-life determinants of illness in Africa: the Drakenstein Child Health Study.* Thorax, 2015. **70**(6): p. 592-594.

7.	Burt, S.A. and K.L. Klump, *The Michigan state university twin registry (MSUTR): An update.* Twin Research and Human Genetics, 2013. **16**(1): p. 344-350.

8.	Klump, K.L. and S.A. Burt, *The Michigan State University Twin Registry (MSUTR): Genetic, environmental and neurobiological influences on behavior across development.* Twin Research and Human Genetics, 2006. **9**(6): p. 971-977.

9.	Burt, S.A. and K.L. Klump, *The Michigan State University Twin Registry (MSUTR): 15 years of twin and family research.* Twin Research and Human Genetics, 2019. **22**(6): p. 741-745.

10.	Mooney, M.A., et al., *Smaller total brain volume but not subcortical structure volume related to common genetic risk for ADHD.* Psychological medicine, 2021. **51**(8): p. 1279-1288.

11.	Mooney, M.A., et al., *Large epigenome-wide association study of childhood ADHD identifies peripheral DNA methylation associated with disease and polygenic risk burden.* Translational Psychiatry, 2020. **10**(1): p. 8.

12.	Nigg, J.T., et al., *The Oregon ADHD-1000: A new longitudinal data resource enriched for clinical cases and multiple levels of analysis.* Developmental cognitive neuroscience, 2023. **60**: p. 101222.

13.	Lawn, W., et al., *The CannTeen Study: Cannabis use disorder, depression, anxiety, and psychotic-like symptoms in adolescent and adult cannabis users and age-matched controls.* Journal of psychopharmacology, 2022. **36**(12): p. 1350-1361.

14.	Kooijman, M.N., et al., *The Generation R Study: design and cohort update 2017.* European journal of epidemiology, 2016. **31**: p. 1243-1264.

15.	Silk, T.J., et al., *Developmental brain trajectories in children with ADHD and controls: a longitudinal neuroimaging study.* BMC psychiatry, 2016. **16**: p. 1-9.

16.	Sciberras, E., et al., *The Children’s Attention Project: a community-based longitudinal study of children with ADHD and non-ADHD controls.* BMC psychiatry, 2013. **13**: p. 1-11.

17.	Saffery, R., et al., *Cohort profile: The peri/post-natal epigenetic twins study.* International journal of epidemiology, 2012. **41**(1): p. 55-61.

18.	Leong, P., et al., *Epigenetic influences on neurodevelopment at 11 years of age: protocol for the longitudinal peri/postnatal epigenetic twins study at 11 years of age (PETS@ 11).* Twin Research and Human Genetics, 2019. **22**(6): p. 446-453.

19.	Min, J.L., et al., *Meffil: efficient normalization and analysis of very large DNA methylation datasets.* Bioinformatics, 2018. **34**(23): p. 3983-3989.

20.	Morin, A.M., et al., *Maternal blood contamination of collected cord blood can be identified using DNA methylation at three CpGs.* Clinical epigenetics, 2017. **9**: p. 1-9.

21.	Aryee, M.J., et al., *Minfi: a flexible and comprehensive Bioconductor package for the analysis of Infinium DNA methylation microarrays.* Bioinformatics, 2014. **30**(10): p. 1363-1369.

22.	Teschendorff, A.E., et al., *A beta-mixture quantile normalization method for correcting probe design bias in Illumina Infinium 450 k DNA methylation data.* Bioinformatics, 2013. **29**(2): p. 189-196.

23.	Leek, J.T., et al., *The sva package for removing batch effects and other unwanted variation in high-throughput experiments.* Bioinformatics, 2012. **28**(6): p. 882-883.

24.	Westra, H.-J., et al., *MixupMapper: correcting sample mix-ups in genome-wide datasets increases power to detect small genetic effects.* Bioinformatics, 2011. **27**(15): p. 2104-2111.

25.	Zheng, S.C., et al., *Identification of differentially methylated cell types in epigenome-wide association studies.* Nature methods, 2018. **15**(12): p. 1059-1066.

26.	Gervin, K., et al., *Systematic evaluation and validation of reference and library selection methods for deconvolution of cord blood DNA methylation data.* Clinical epigenetics, 2019. **11**: p. 1-15.

27.	Dieckmann, L., et al., *Reliability of a novel approach for reference-based cell type estimation in human placental DNA methylation studies.* Cellular and Molecular Life Sciences, 2022. **79**(2): p. 115.

28.	Du, P., W.A. Kibbe, and S.M. Lin, *lumi: a pipeline for processing Illumina microarray.* Bioinformatics, 2008. **24**(13): p. 1547-1548.

29.	Pidsley, R., et al., *A data-driven approach to preprocessing Illumina 450K methylation array data.* BMC genomics, 2013. **14**: p. 1-10.

30.	BeadChip, M., *Identification of polymorphic and off-target probe binding sites on the Illumina Infinium.*

31.	Ollikainen, M., et al., *DNA methylation analysis of multiple tissues from newborn twins reveals both genetic and intrauterine components to variation in the human neonatal epigenome.* Human molecular genetics, 2010. **19**(21): p. 4176-4188.

32.	Maksimovic, J., et al., *Removing unwanted variation in a differential methylation analysis of Illumina HumanMethylation450 array data.* Nucleic acids research, 2015. **43**(16): p. e106-e106.

33.	Bakulski, K.M., et al., *DNA methylation of cord blood cell types: applications for mixed cell birth studies.* Epigenetics, 2016. **11**(5): p. 354-362.

34.	Sharp, T.H., et al., *Population neuroimaging: generation of a comprehensive data resource within the ALSPAC pregnancy and birth cohort.* Wellcome Open Research, 2020. **5**.

35.	Shi, F., et al., *Infant brain atlases from neonates to 1-and 2-year-olds.* PloS one, 2011. **6**(4): p. e18746.

36.	Tzourio-Mazoyer, N., et al., *Automated anatomical labeling of activations in SPM using a macroscopic anatomical parcellation of the MNI MRI single-subject brain.* Neuroimage, 2002. **15**(1): p. 273-289.

37.	Feczko, E., et al., *Adolescent Brain Cognitive Development (ABCD) community MRI collection and utilities.* BioRxiv, 2021: p. 2021.07. 09.451638.

38.	Dale, A.M., B. Fischl, and M.I. Sereno, *Cortical surface-based analysis: I. Segmentation and surface reconstruction.* Neuroimage, 1999. **9**(2): p. 179-194.

39.	Avants, B.B., et al., *A reproducible evaluation of ANTs similarity metric performance in brain image registration.* Neuroimage, 2011. **54**(3): p. 2033-2044.

40.	Dubos, J., et al. *Dmriprep: open-source diffusion MRI quality control framework with graphical user interface*. in *Medical Imaging 2023: Image Processing*. 2023. SPIE.

41.	Jenkinson, M., et al., *Fsl.* Neuroimage, 2012. **62**(2): p. 782-790.

42.	Houghton, A., et al., *BIBSnet (3.2.0). .* Zenodo, 2024.

43.	Goncalves, M., et al. *NiBabies: a robust preprocessing workflow tailored for neonate and infant MRI*. in *27th Annual Meeting of the Organization for Human Brain Mapping*. 2021.

44.	Mehta, K., et al., *XCP-D: A Robust Pipeline for the post-processing of fMRI data.* bioRxiv, 2023.

45.	Hagler Jr, D.J., et al., *Image processing and analysis methods for the Adolescent Brain Cognitive Development Study.* Neuroimage, 2019. **202**: p. 116091.

46.	Jones, S.A., et al., *Attention‐deficit/hyperactivity disorder and white matter microstructure: The importance of dimensional analyses and sex differences.* JCPP advances, 2022. **2**(4): p. e12109.

47.	Skumlien, M., et al., *Neural responses to reward anticipation and feedback in adult and adolescent cannabis users and controls.* Neuropsychopharmacology, 2022. **47**(11): p. 1976-1983.

48.	Acosta, H., et al., *Partial support for an interaction between a polygenic risk score for major depressive disorder and prenatal maternal depressive symptoms on infant right amygdalar volumes.* Cerebral Cortex, 2020. **30**(12): p. 6121-6134.

49.	White, T., et al., *Paediatric population neuroimaging and the Generation R Study: the second wave.* European Journal of Epidemiology, 2018. **33**: p. 99-125.

50.	Blumenthal, J.D., et al., *Motion artifact in magnetic resonance imaging: implications for automated analysis.* Neuroimage, 2002. **16**(1): p. 89-92.

51.	Cherel, M., et al. *Automatic tissue segmentation of neonate brain MR Images with subject-specific atlases*. in *Medical Imaging 2015: Image Processing*. 2015. SPIE.

52.	Gilmore, J.H., et al., *Regional gray matter growth, sexual dimorphism, and cerebral asymmetry in the neonatal brain.* Journal of Neuroscience, 2007. **27**(6): p. 1255-1260.
